# Skin Cancer Classification Using Explainable Artificial Intelligence With an Ensemble Model and Rigorous Leakage Free Validation

**DOI:** 10.64898/2026.09.02.26362011

**Authors:** Mehmet Tarık Baran, Ömer Karakoyun

**Author notes:** Corresponding author: Mehmet Tarık Baran. Ömer Karakoyun.

## Abstract

**Background:** Reliable melanoma classification requires models that capture both local dermoscopic morphology and broader contextual patterns while maintaining auditable, leakageaware internal validation.

**Objectives:** To develop and internally validate an EfficientNetB0–Swin Transformer Tiny ensemble for classifying histopathologically verified dermoscopic images as benign melanocytic lesions or malignant melanoma.

**Methods:** This retrospective diagnostic model-development and internal validation study screened 552,869 ISIC Archive records; filtering and dermatologist review yielded 1,199 uniquepatient and unique lesion images (578 benign and 621 malignant). Images were the predictors and histopathology was the reference. ImageNet pretrained EfficientNetB0 and Swin-T features were fused. Patient independent five fold validation used weighted sampling, mixup, label smoothing, AdamW, early stopping, and five view test time augmentation.

**Results:** Mean accuracy was 0.89325 ± 0.03179, mean receiver operating characteristic area under the curve (ROC-AUC) was 0.96348 ± 0.01695, and mean support weighted F1-score was 0.89300 ± 0.03220. The fold level 95% confidence intervals were 0.8538–0.9327 for accuracy and 0.9424–0.9845 for ROC-AUC. Pooled counts were 526 true negatives, 52 false positives, 76 false negatives, and 545 true positives, yielding 87.76% sensitivity and 91.00% specificity. Qualitative Grad-CAM review showed peripheral artifact activation in two false positives and lesion centered activation in two correctly classified cases; these observations were not systematically scored.

**Limitations:** The validation folds were also used for early stopping and checkpoint selection. Device stratified analysis, systematic interpretability scoring, calibration, and independent external validation were unavailable.

**Conclusions:** The ensemble showed high internal discrimination and is intended only as a clinician facing adjunct. The error audit workflow enables targeted retrospective review, but external validation is required before clinical use or generalizability claims. The study was not registered.

## 1 Introduction

The present study addresses binary classification of histopathologically verified dermoscopic images into benign melanocytic lesions and malignant melanoma. The modeling cohort was derived from the publicly accessible International Skin Imaging Collaboration (ISIC) Archive using explicit image type, diagnostic reference, lesion, image manipulation, and metadata filters. The central methodological challenge was to construct a model that could represent both local image patterns and broader contextual relationships while maintaining stable performance across stratified validation subsets.

Cutaneous melanoma represents a substantial medical burden. Global estimates for 2020 reported approximately 325,000 new cases and 57,000 deaths, with marked increases in both incidence and mortality projected by 2040 [1]. This burden reinforces the need for reliable pathways that support early recognition, appropriate referral, and timely histopathologic assessment.

From a medical perspective, an automated dermoscopic classifier should be positioned as a clinical decision-support and quality assurance tool rather than as a replacement for dermatologic examination or histopathology. False negative classification of melanoma is the most consequential model error because it may falsely lower suspicion for a malignant lesion, whereas false positive classification may increase referrals, biopsies, patient anxiety, and resource use. Accordingly, clinically meaningful evaluation requires not only discrimination and accuracy but also threshold specific sensitivity and specificity, probability calibration, transparent error review, and interpretable visual evidence.

The target population is patients with melanocytic lesions undergoing dermoscopic assessment in specialist dermatology or referral settings. Intended users are dermatologists and other appropriately trained clinicians. The model is intended to support lesion prioritization, closer examination, referral, surveillance, or biopsy decisions as a second reading aid; it is not intended for direct patient self use or autonomous diagnosis.

To this end, a heterogeneous feature level ensemble was developed from two ImageNet-1K-pretrained backbones. EfficientNetB0 served as the convolutional branch and produced a 1,280-dimensional feature vector [2]. Swin Transformer Tiny served as the transformer branch and produced a 768-dimensional embedding [3]. The two representations were concatenated and classified jointly, allowing the final decision layer to use information from both branches.

The training protocol combined several regularization and robustness mechanisms: geometric and photometric image augmentation, coarse dropout, weighted random sampling, mixup, label smoothing, AdamW optimization, early stopping, and five view test time augmentation (TTA). Internal performance was assessed by stratified five-fold cross-validation. Post hoc interpretability and representation analyses used gradient weighted class activation mapping (Grad-CAM) and Uniform Manifold Approximation and Projection (UMAP). These analyses were interpreted qualitatively and were not treated as independent performance measures.

The objective was to develop and internally validate the proposed EfficientNetB0–Swin-T ensemble for dermoscopic melanoma classification and to quantify its fold wise discrimination and classification performance. The primary monitored validation metric was ROC-AUC. Accuracy, precision, recall, and F1-score were additional performance measures.

### 1.1 Relation to prior work and study positioning

Deep ensembles, CNN–Transformer hybrids, and explainable hybrid systems have previously been investigated for dermoscopic image classification. Harangi fused the outputs of multiple CNN architectures and reported that the ensemble outperformed the individual networks [4]. Guergueb and Akhloufi subsequently grouped deep-model predictions for automated melanoma detection [5], while Chiu et al. extended heterogeneous ensemble learning to combinations of CNNs and vision transformers [6]. Explainable hybrid systems have also integrated vision-transformer, CNN, and Xception representations with Grad-CAM across HAM10000 and ISIC data [7], and modified EfficientNetB0 systems have been optimized across multiple dermoscopic benchmarks with explicit consideration of computational cost [8]. Other related approaches include ensembles of fine tuned CNNs, end to end CNN–Transformer models, an EfficientNet–Swin Transformer architecture with Grad-CAM, and more recent explainable hybrid melanoma frameworks [9, 10, 11, 12]. Consequently, an absolute claim that no comparable algorithm exists in the literature would not be supportable.

The contribution of the present study is instead the specific integration of ImageNet-pretrained EfficientNetB0 and Swin-T feature vectors within a histopathology-filtered, near balanced binary ISIC cohort, together with weighted sampling, mixup, label smoothing, five view TTA, stratified cross validation, and an image level error audit workflow. Whether this complete configuration provides an advantage over previously reported systems must ultimately be established through same cohort baseline comparisons and ablation experiments.

Dermatology AI may perform unequally across skin tones, demographic groups, disease subtypes, devices, and care settings [13, 14]. Skin tone, race, ethnicity, socioeconomic status, and device metadata were unavailable for the present cohort; corresponding health-inequality and fairness effects could therefore not be evaluated.

## 2 Materials and Methods

### 2.1 Study design

This was a computational model development and internal validation study based on a retrospectively filtered subset of the ISIC Archive. The workflow comprised six stages: (1) archive screening and cohort construction, (2) image preprocessing and augmentation, (3) construction of the heterogeneous ensemble, (4) stratified five fold partitioning, (5) fold-specific training with mixup and TTA-based validation, and (6) post hoc performance and interpretability analyses.

The manuscript was prepared in accordance with the TRIPOD+AI guidance for prediction model reporting and with reference to the CLAIM 2024 recommendations for artificial intelligence studies in medical imaging [15, 16]. These frameworks guided transparent reporting of the data source, eligibility criteria, reference standard, partitioning strategy, model specification, performance measures, interpretability analyses, and limitations; they do not imply prospective or external validation.

The unit of analysis was a unique patient lesion pair. The final cohort contained 1,199 unique patients, 1,199 unique lesions, and exactly one image per patient and lesion. Thus, no patient or lesion could occur in more than one cross validation fold. The acquisition period was not available, and no independent external test cohort was used. No prospective treatment allocation or follow up was involved; treatment information and a prediction time horizon were therefore not applicable to this diagnostic image classification study.

### 2.2 Data source and contributor metadata

Images were obtained from the publicly accessible ISIC Archive and stored on the local workstation in a folder structure compatible with image folder classification. Machine-specific absolute paths were not retained in the manuscript. The source records represented multiple contributors, but contributor specific image counts were unavailable and no contributor stratified analysis was performed.

The same archive derived cohort supplied both model development and internal validation folds. ISIC was selected because it provides dermoscopic images with diagnostic metadata and histopathologic reference labels, but it is not a consecutive clinical cohort and may not represent the intended specialist care population. Image acquisition and participant accrual dates, and the number and locations of contributing centers represented in the final cohort, were unavailable.

The ISIC Archive assigns licenses at the image contributor level; consequently, the exact license and required attribution for each image in the final cohort must be preserved in the study manifest.

### 2.3 Cohort construction

At the time of cohort construction, 552,869 ISIC images were screened. Restriction to Image Type = dermoscopic yielded 124,811 images. Restriction to Type of Diagnosis = histopathology yielded 42,068 images with histopathologic reference diagnoses. Indeterminate records were excluded before the benign and malignant selection branches were finalized.

For the benign branch, the metadata filters required *Melanocytic* to be true and *Image Manipulation* to be instrument only. Accepted *Lesion Diagnosis* values were BENIGN, BENİGN MELANOCYTİC PROLİFERATİONS, and Nevus. This query produced 2,494 eligible benign images; 578 images were retained after metadata verification and dermatologist-led quality review.

For the malignant branch, the metadata filters likewise required Melanocytic to be true and image manipulation to be instrument only. Accepted *Lesion Diagnosis* values were MALIGNANT and MALIGNANT MELANOCYTIC PROLIFERATIONS (MELANOMA). This query produced 973 eligible malignant images; 621 images were retained after metadata verification and dermatologist-led quality review.

The selection process is summarized in Figure 1.

**Figure 1.**
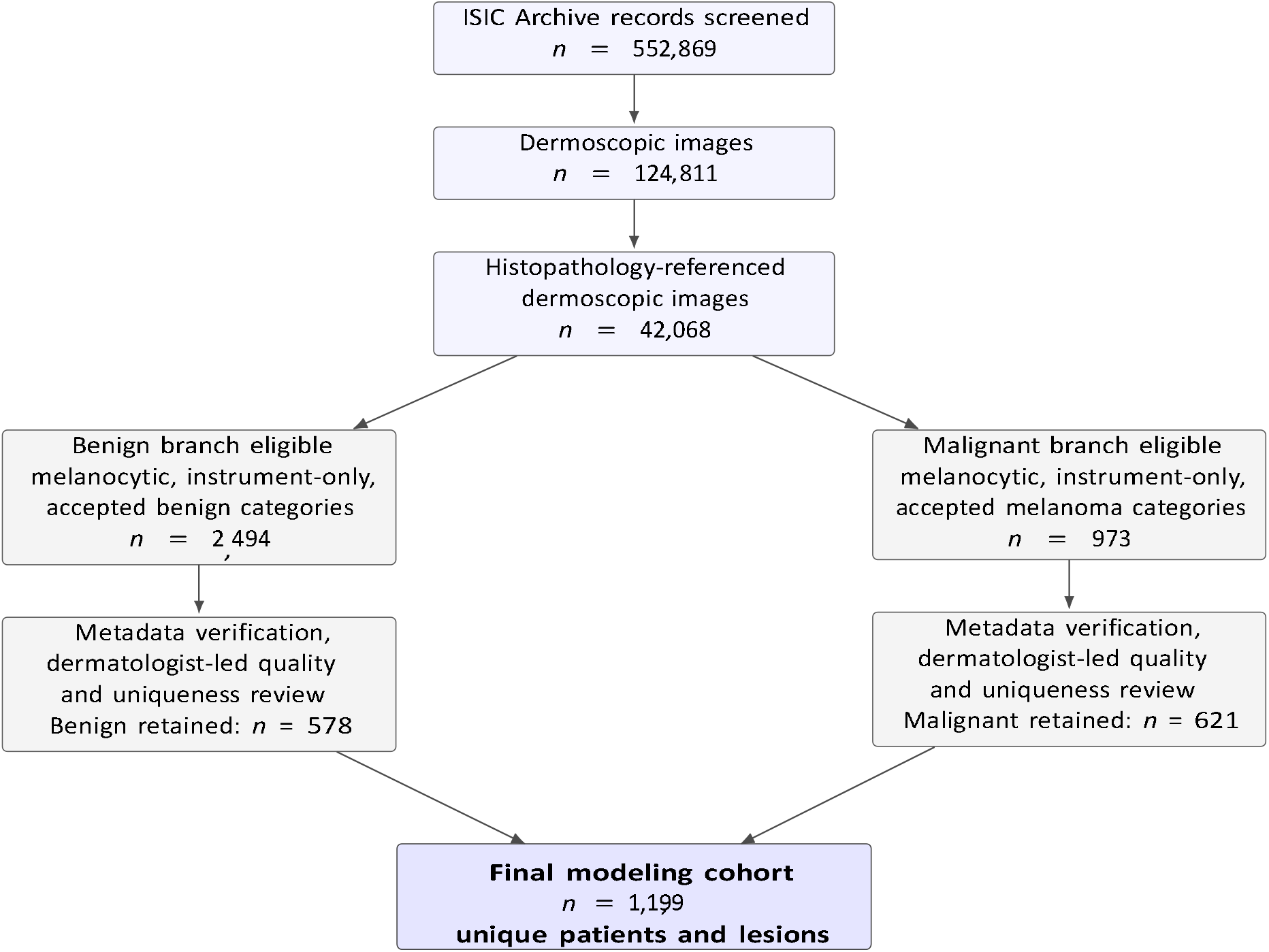
Flow diagram of ISIC Archive screening and cohort construction. Indeterminate diagnostic records were excluded before the benign and malignant branches were finalized. Reductions from the eligible branches to the retained subsets resulted from metadata verification and dermatologist led image quality and uniqueness review; reason-specific exclusion counts were not recorded.

Eight fields were mandatory at cohort selection: image type, diagnostic-reference type, melanocytic status, lesion diagnosis, image-manipulation status, sex, approximate age, and anatomic site. All 2,494 benign-eligible and 973 malignant-eligible records contained these fields, and all final images retained complete values. The reduction to the final cohort was therefore attributable to manual image selection and quality review rather than missingness in these eight fields.

Image quality, visible artifacts, blur, and focus were reviewed manually by a dermatologist. Peripheral dark dermatoscope borders and related acquisition artifacts were qualitatively frequent in the eligible image pool before final selection. ISIC image identifiers and manual review were used to exclude exact or visually near duplicate records and multiple views of the same lesion. Only one image per patient and lesion was retained. Overall, 1,916 benign-eligible images (76.8%) and 352 malignanteligible images (36.2%) were excluded during final selection; artifact prevalence and reason-specific exclusion counts were not recorded and therefore could not be quantified.

### 2.4 Final dataset and image organization

The final dataset contained 1,199 dermoscopic JPEG images with histopathologic reference diagnoses, representing 1,199 unique patients and 1,199 unique lesions. Images were assigned to one of two folders and corresponding target classes:

- Benign : benign melanocytic lesion;
- Malignant : malignant melanoma.

The benign class contained 578 images (48.2%), and the malignant class contained 621 images (51.8%), for a total of 1,199 images. Each image corresponded to a different patient and lesion. Images were read as three-channel RGB images. Original image dimensions varied. All images were resized to 224 x 224 pixels before model input. For each cross validation iteration, approximately 80% of samples formed the training subset and the complementary 20% formed the validation subset. Stratification preserved the available class proportions in each fold.

### 2.5 Clinical and imaging characteristics

The supplied aggregate metadata included age group, sex, anatomic site, and malignant lesion subtype. Age, sex, and anatomic site distributions are summarized together in Table 1. Mean age was approximately 48 years in the benign group and 64 years in the malignant group. The malignant group was more frequently male than the benign group. Palms and soles were the most frequent site among malignant images, whereas the lower extremity was the most frequent site among benign images. All 1,199 images were allocated to an age group, sex category, and anatomic site category.

**Table 1.** Clinical and imaging characteristics of the final cohort. Percentages use the corresponding class denominator.

| Characteristic/category | Benign ($n = 578$) | | Malignant ($n = 621$) | |
| --- | --- | --- | --- | --- |
| | $n$ | % | $n$ | % |
| *Age group <* |  |  |  |  |
| 30 years | 67 | 11.6 | 11 | 1.8 |
| 30–44 years | 154 | 26.6 | 45 | 7.2 |
| 45–59 years | 167 | 28.9 | 127 | 20.5 |
| 60–75 years | 164 | 28.4 | 279 | 44.9 |
| > 75 years | 26 | 4.5 | 159 | 25.6 |
| Unallocated | 0 | 0.0 | 0 | 0.0 |

| Approximate mean, years | 48 |  | 64 |  |
| --- | --- | --- | --- | --- |
| *Sex* |  |  |  |  |
| Male | 208 | 36.0 | 297 | 47.8 |
| Female | 370 | 64.0 | 324 | 52.2 |
| *Anatomic site* Palms/soles | 187 | 32.4 | 360 | 58.0 |
| Lower extremity | 227 | 39.3 | 103 | 16.6 |
| Upper extremity | 84 | 14.5 | 111 | 17.9 |
| Oral/genital | 70 | 12.1 | 33 | 5.3 |
| Head/neck | 10 | 1.7 | 14 | 2.3 |

Within the malignant class, the supplied subtype proportions were 48.3% melanoma in situ, 42.3% invasive melanoma, 8.2% melanoma not otherwise specified, and 1.2% metastatic melanoma. These percentages correspond to approximate counts of 300, 263, 51, and 7 images, respectively, after rounding. Capture year, exact acquisition center per image, dermatoscope manufacturer, and device model were not available for analysis.

### 2.6 Outcome, predictors, assessors, study size, and missing data

The diagnostic outcome was malignant melanoma versus a benign melanocytic lesion. Histopathology recorded in the ISIC Archive was the reference standard and no prognostic time horizon applied. All included images required a histopathologic reference; however, the original pathology procedures, timing relative to dermoscopy, assessor qualifications and demographic characteristics, and blinding procedures were unavailable. Consistency of outcome assessment across sociodemographic groups could not be evaluated.

The sole model predictor was the three channel RGB dermoscopic image, represented through automatically learned pixel derived features. Age, sex, anatomic site, contributor, and melanomasubtype metadata were not model inputs. A dermatologist performed image quality and uniqueness review, but the reviewer’s experience details, demographic characteristics, and blinding to the reference diagnosis were not recorded.

No formal sample-size or precision calculation preceded this secondary analysis. Study size was determined by archive eligibility, completeness of the eight mandatory fields, image quality, uniqueness, and the one patient/one lesion rule. The mandatory fields were complete in all branch eligible and retained records; no imputation was performed. Records excluded during manual review were omitted rather than imputed, and reason-specific exclusion counts were unavailable.

### 2.7 Implementation and reproducibility

The analysis pipeline was implemented in Python 3.10 or later with PyTorch 2.0.1 and torchvision 0.15.2. Image loading and preprocessing used Pillow 10.0.0 and Albumentations 1.3.1. Data management and evaluation used NumPy 1.24.3, pandas 2.0.3, and scikit-learn 1.3.0; visualization and post hoc analyses used Matplotlib 3.7.2, seaborn 0.12.2, umap-learn 0.5.3, and pytorch-gradcam 1.4.6. The recorded CUDA runtime was 11.8. The software stack, pretrained initialization, deterministic settings, and device configuration were reported in line with comparable dermoscopic AI studies and current medical-imaging prediction-model guidance [9, 10, 7, 8, 15, 16].

Execution was configured for a single compute device. The code selected a CUDA enabled NVIDIA GPU when available and otherwise used the CPU. The exact operating system, processor, GPU model, GPU memory, and system memory were not preserved; consequently, hardware specific training time, memory demand, inference latency, and energy consumption cannot be reproduced from the available records. No distributed training or multi device aggregation was specified.

EfficientNetB0 and Swin-T were initialized from torchvision IMAGENET1K_V1 weights; the recorded weight files were efficientnet_b0_rwightman-7f5810bc.pth and swin_t-704ceda3.pth, respectively. A new ensemble was instantiated and optimized independently within each cross validation fold. The Python random, NumPy, and PyTorch random number generators were initialized with seed 42. Deterministic cuDNN execution was requested with torch.backends.cudnn.deterministic= True, and cuDNN benchmarking was disabled. The training DataLoader used a batch size of 32 and the PyTorch default of zero worker processes because num_workers was not overridden. Mixed precision training, gradient accumulation, and a separate hyperparameter search framework were not documented in the supplied implementation.

### 2.8 Image preprocessing and augmentation

Images were opened with Pillow, converted to RGB, and represented as NumPy arrays before Albumentations transformations. The resulting tensor shape was 3 x 224 x 224.

#### 2.8.1 Training transformations

The training pipeline applied the following operations in sequence:

1. resize to 224 x 224 pixels;
2. horizontal flip with probability 0.5;
3. random 90^⍰^ rotation with probability 0.5;
4. color jitter with brightness, contrast, saturation, and hue magnitudes of 0.2 and application probability 0.5;
5. coarse dropout with at most eight masked regions, maximum region height of 16 pixels, maximum region width of 16 pixels, and application probability 0.5;
6. channel-wise normalization with mean (0.5,0.5,0.5) and standard deviation (0.5,0.5,0.5), followed by conversion to a PyTorch tensor. For channel *c*, normalization was performed as

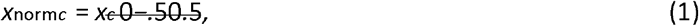

mapping input intensities from [0,1] to [−1,1]. Horizontal flipping and right-angle rotation introduced geometric variation; color jitter introduced photometric variation; and coarse dropout masked small image regions during training.

#### 2.8.2 Validation transformations

The base validation transform was deterministic and consisted only of resizing to 224 × 224 pixels, applying the same channel-wise normalization, and converting the image to a tensor. No stochastic operation was used in this base transform. Separately, the TTA procedure generated stochastic views at inference as described below.

#### 2.8.3 Dataset implementation

The custom dataset stored image-path and integer-label pairs. For a requested index, the image was opened, converted to RGB, transformed, and returned with its label. Thus, the data path was path → PIL RGB image → NumPy array → Albumentations → normalized tensor.

### 2.9 Heterogeneous ensemble architecture

The ensemble was selected to balance four design priorities: predictive accuracy, complementary local and global image representation, suitability for heterogeneous dermoscopic appearances and contributor sources, and computational economy. EfficientNetB0 provides parameter-efficient convolutional feature extraction, whereas Swin-T contributes hierarchical shifted-window attention for broader contextual relationships. The B0 and Tiny variants were deliberately preferred over larger backbone variants to maximize expected predictive utility per unit of computation while limiting memory, runtime, and anticipated energy demand. This choice was informed by prior evidence that ensemble and hybrid fusion can improve skin-lesion classification and that EfficientNet-family architectures can balance accuracy with computational cost [2, 3, 4, 6, 7, 8]. However, floating-point operation counts, inference latency, electrical energy consumption, and carbon emissions were not measured; reduced energy use was therefore a design objective rather than a demonstrated study outcome.

#### 2.9.1 EfficientNetB0 branch

The EfficientNetB0 model was initialized with ImageNet-1K pretrained weights. Its original classifier was replaced by an identity mapping, yielding a 1,280-dimensional feature vector for each image:

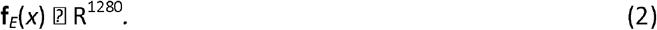

The supplied implementation reported 5,330,564 parameters for the unmodified EfficientNetB0 model. The branch comprised the convolutional stem, mobile inverted bottleneck blocks, a final convolutional representation, and global average pooling.

#### 2.9.2 Swin Transformer Tiny branch

Swin-T was also initialized with ImageNet-1K pretrained weights. Its classification head was replaced by an identity mapping, yielding a 768-dimensional embedding:

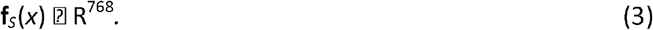

The supplied implementation reported 28,288,354 parameters for the unmodified Swin-T model. Images were represented using 4 × 4 patch embedding, hierarchical feature stages with dimensions 96, 192, 384, and 768, and shifted-window attention with a 7 × 7 window.

#### 2.9.3 Feature fusion and classification head

The two backbone outputs were concatenated along the feature dimension:

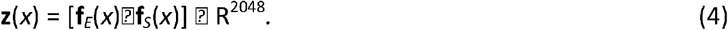

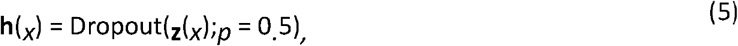

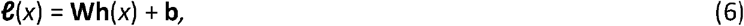

where **W**⍰ *R*^2×2048^ and **b** ⍰ *R*^2^. Class probabilities were computed using

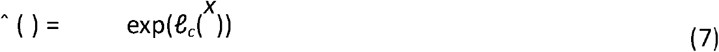

The fused representation was passed through dropout with *p* = 0.5 and a linear layer with two outputs:

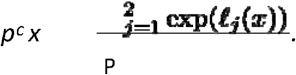

The linear head contained 4,098 parameters. The supplied notes stated a total of 33,623,016 parameter by summing the two baseline-model counts and the new linear head. Because the original 1,000-class heads were replaced by identity mappings, the exact instantiated and trainable parameter counts must be confirmed from the executed model rather than inferred from that baseline sum.

### 2.10 Stratified five-fold cross-validation

Stratified K-fold cross-validation was configured with five splits, shuffling enabled, and random_ state=4 For dataset

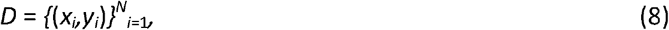

the split procedure aimed to preserve the class distribution in each fold:

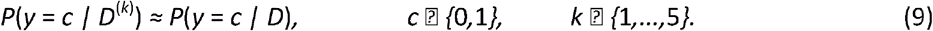

For each fold, the model was trained independently on four partitions and validated on the remaining partition. Training and validation indices did not overlap within a fold.

The split was implemented over image samples. Because cohort construction retained exactly one image for each unique patient and lesion, image-level partitioning was also patient- and lesionindependent. Exact duplicate records were excluded by ISIC identifier and visually near-duplicate or multi-view cases were excluded during manual review; consequently, no patient or lesion was represented in more than one fold.

Sampling weights and stochastic training transformations were calculated or applied only within the training portion of each fold, and validation images were not used for optimizer updates. The held-out fold was nevertheless used for early stopping and checkpoint selection as well as for reporting fold performance; it should therefore be interpreted as an internal validation fold rather than as a completely untouched test set.

### 2.11 Class balancing by weighted random sampling

Within each training fold, inverse-frequency sample weights were computed from the class counts. If *n*_*c*_ denotes the number of training images in class *c*, the implemented per-sample weight was proportional to

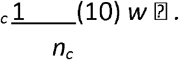

A weighted random sampler then drew the same number of samples as the training-fold size, with replacement. The relative sampling weight between any two classes was therefore inversely proportional to their observed frequencies in that training fold. Samples were delivered in batches of 32.

Weighted sampling addressed diagnostic-class imbalance only. It was not a demographic fairness intervention, and no post hoc probability recalibration was performed.

### 2.12 Training protocol

#### 2.12.1 Mixup

For every training batch, mixup generated convex combinations of randomly paired samples. A mixing coefficient was sampled as

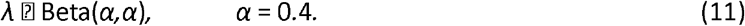

Given a random batch permutation *π*, mixed inputs were

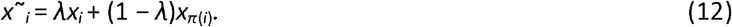

The batch loss combined the losses for the two associated labels:

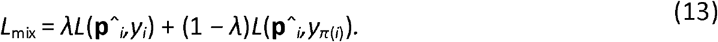

#### 2.12.2 Label-smoothed cross-entropy

The criterion was cross-entropy with label smoothing *ε* = 0.1. For *C* = 2 classes, the smoothed target was

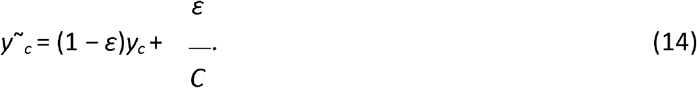

Thus, a one-hot target [1,0] became [0.95,0.05]. The loss was

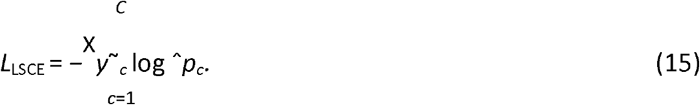

During mixup training, this criterion was evaluated for each of the paired hard labels and combined using *λ*.

#### 2.12.3 Optimization

All model parameters were optimized using AdamW with learning rate 10^−4^, *β*_1_ = 0.9, *β*_2_ = 0.999, numerical-stability constant ⍰ = 10^−8^, and decoupled weight decay 10^−4^. No backbone layer was frozen; both pretrained branches and the fusion classifier were fine-tuned jointly. No learning-rate scheduler was specified in the supplied protocol.

#### 2.12.4 Early stopping and checkpointing

Validation ROC-AUC was monitored after each epoch. If ROC-AUC improved, the current model state was saved as the best checkpoint for that fold and the waiting counter was reset. Training stopped after six consecutive epochs without improvement. The maximum training horizon was 30 epochs. The checkpoint with the highest validation ROC-AUC was retained for each fold. Among the five fold-specific checkpoints, Fold 4 had the highest reported validation ROC-AUC. The implementation copied the highest-AUC fold checkpoint to the global model file best_isic_model.pth; it did not contain a subsequent step that retrained a final model on the complete dataset.

#### 2.12.3 Test-time augmentation

TTA generated five independently augmented views of each validation image using the complete stochastic training augmentation pipeline, including horizontal flipping, random 90^⍰^ rotation, color jitter, and coarse dropout. Softmax probabilities from the five forward passes were averaged:

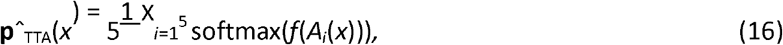

where *A*_*i*_ denotes the *i*th stochastic transformation and *f* is the ensemble. The predicted class was selected by the maximum mean probability, equivalent to a binary threshold of 0.5 for the malignant-class probability. For the alphabetically ordered image folders, benign corresponded to class 0 and malignant to class 1; the class-1 probability was used for binary ROC-AUC calculation.

This threshold was not clinically optimized.

### 2.14 Performance outcomes

For the medical interpretation of the binary task, malignant melanoma was treated as the positive class and benign lesions as the negative class. True positives (TP) were malignant images classified as malignant; false negatives (FN) were malignant images classified as benign; false positives (FP) were benign images classified as malignant; and true negatives (TN) were benign images classified as benign.

Threshold-dependent metrics were defined as

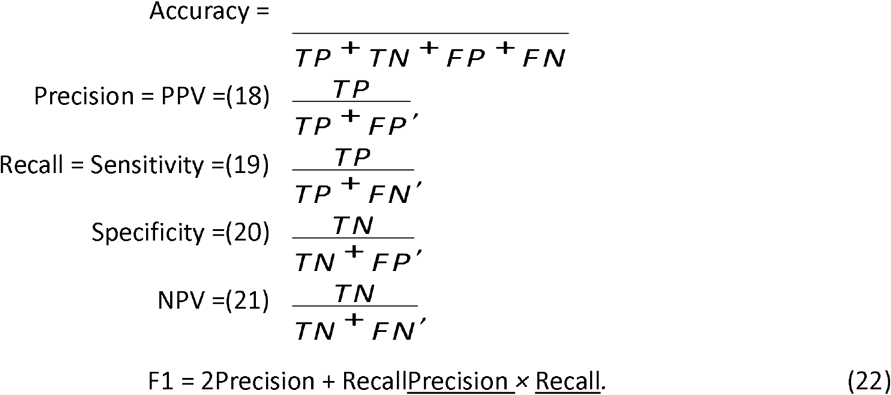

The ROC curve related the true-positive rate to the false-positive rate across classification thresholds, and ROC-AUC summarized discrimination independent of one fixed threshold. The supplied global plots reported a pooled ROC-AUC of 0.962 and a pooled precision–recall AUC (PR-AUC) of 0.969 (Figure 2c–d).

**Figure 2.**
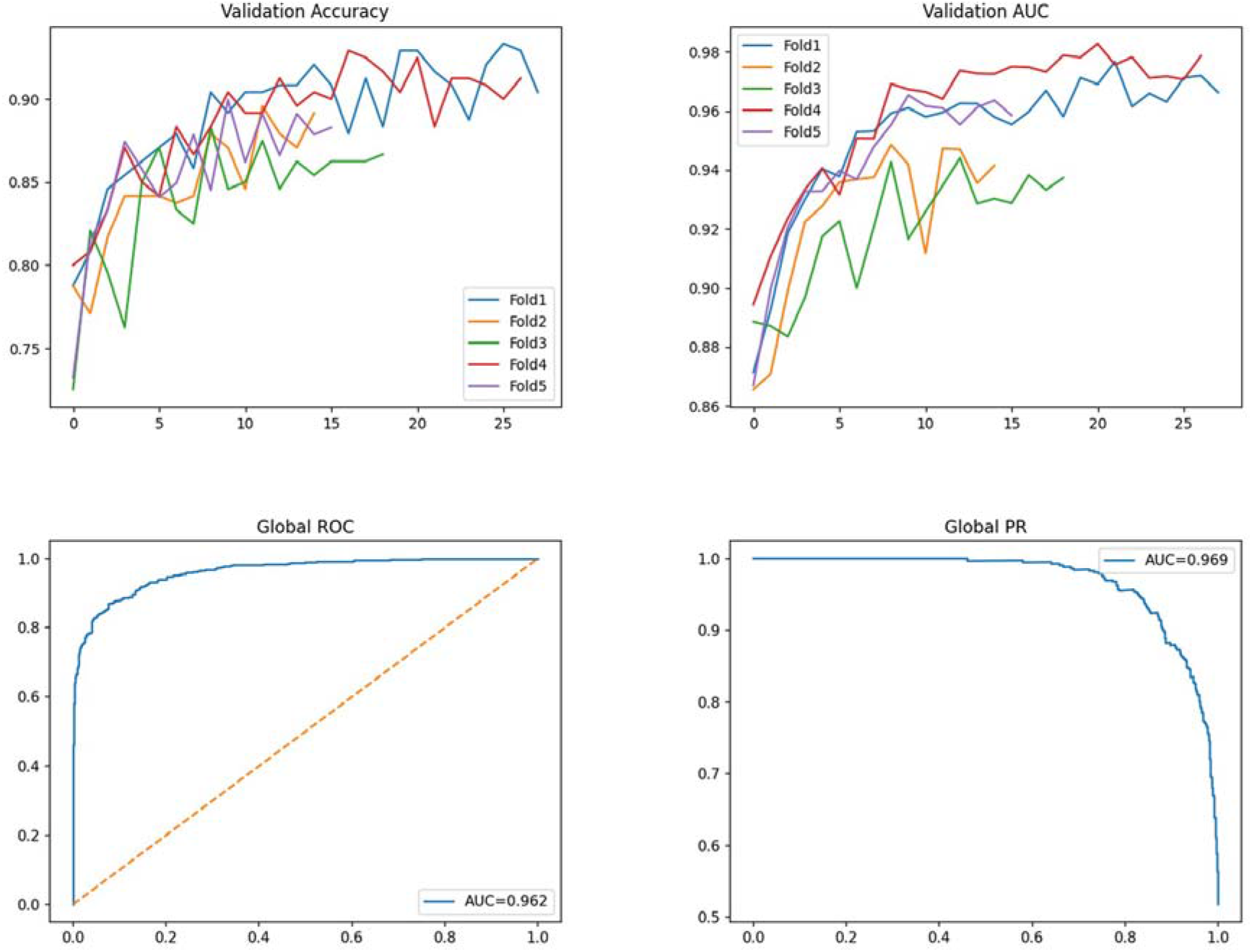
Global performance plots. (a) Fold-specific validation accuracy trajectories; (b) fold-specific validation receiver operating characteristic area under the curve (ROC-AUC) trajectories; (c) global receiver operating characteristic curve (AUC = 0.962); and (d) global precision–recall curve (AUC = 0.969). Different trajectory lengths reflect fold-specific stopping times.

The reported precision, recall, and F1-score values were calculated using scikit-learn’s supportweighted averaging. For a class-specific metric *M*_*c*_, the reported aggregate was

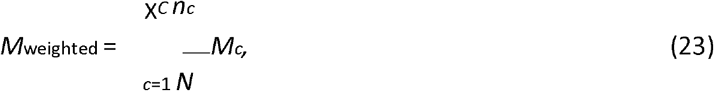

where *n*_*c*_ is the number of validation samples in class *c* and *N* is the validation-fold size.

### 2.15 Statistical analysis

Fold-level performance was summarized using the arithmetic mean, sample standard deviation, minimum, maximum, and range. All summary values in this manuscript were recalculated directly from the five supplied fold-level rows. Descriptive 95% confidence intervals were calculated using

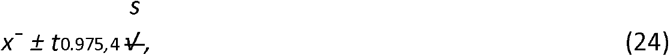

where 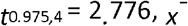 is the fold mean, and *s* is the sample standard deviation of the five fold values. The coefficient of variation was computed as 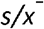.

Class differences in sex and the grouped anatomic-site categories were evaluated using Pearson’s chi-square test. Effect size was summarized with Cramér’s *V* . For sex, the odds ratio (OR) for male sex in malignant versus benign records was calculated with a log-scale 95% confidence interval. For palms/soles location, a prevalence ratio (PR) and log-scale 95% confidence interval were calculated. Age-group distributions and approximate mean ages were summarized descriptively. All tests were two-sided, and *p <* 0.05 was considered statistically significant. These cohort-composition analyses describe selection differences and do not measure model performance within demographic subgroups.

Calibration, decision-curve analysis, and other clinical-utility measures were not performed. Performance was not evaluated within age, sex, anatomic-site, skin-tone, race, ethnicity, device, contributor, or melanoma-subtype groups. Contributor-level clustering and between-center heterogeneity were not modeled because contributor-specific counts were unavailable. No model updating or recalibration followed internal validation.

### 2.16 Interpretability and representation analyses

#### 2.16.1 Grad-CAM

Grad-CAM was configured for the last feature block of the EfficientNetB0 branch [17]. For class score *Y*^*c*^

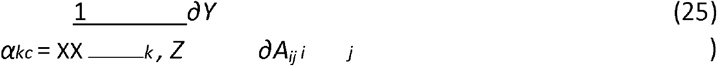

where *Z* is the number of spatial locations. The class-activation map was then

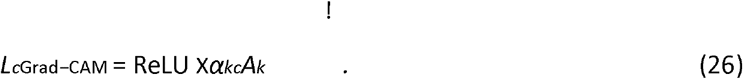

and feature map *A*^*k*^, channel importance weights were calculated as

The normalized heat map was overlaid on the original dermoscopic image and saved beside the original image. The reporting pipeline was configured to generate Grad-CAM visualizations for both correctly and incorrectly classified validation images. Two Fold 1 false-positive cases and two correctly classified Fold 1 cases were available for qualitative review. Review considered whether activation overlapped the lesion, lesion margins and perilesional skin, or noncutaneous peripheral artifacts. A systematic expert assessment or quantitative localization score was unavailable.

#### 2.16.2 UMAP

For representation analysis, 1,280-dimensional EfficientNetB0 features and 768-dimensional Swin-T embeddings were concatenated to form the same 2,048-dimensional representation used by the classifier. The analyzed subset comprised all recorded misclassifications followed by up to the first 200 correctly classified images. UMAP was instantiated without explicitly overriding its hyperparameters and projected these vectors into two dimensions for class-wise visualization [18]. The projection was interpreted qualitatively; no quantitative cluster-separation statistic was calculated.

### 2.17 Error-audit and reporting workflow

For every labeled validation image, the predicted class was compared programmatically with the histopathologic reference label. All discordant cases were assigned to the misclassification collection, while concordant cases were assigned to a separate correctly classified collection. The reporting workflow preserved the fold identifier, image path, reference class, predicted class, and class-1 probability; copied the corresponding images into audit directories; and was configured to generate paired original-image and Grad-CAM visualizations. Separate HTML reports and comma-separated summary files were generated for correct and incorrect predictions.

This procedure can identify every error relative to a known reference label within the retrospective validation folds and makes those errors available for systematic review. It does not enable the model to recognize autonomously that a prediction is wrong when the reference diagnosis is unknown in prospective use.

## 3 Results

### 3.1 Cohort assembly

Of the 552,869 ISIC images screened, 124,811 met the dermoscopic image-type criterion and 42,068 had a histopathologic diagnostic reference. The benign selection branch yielded 2,494 records meeting all eight mandatory fields; 578 were retained after dermatologist-led image-quality and uniqueness review. The malignant melanoma branch yielded 973 records meeting the mandatory fields; 621 were retained. The corresponding exclusion proportions were 76.8% and 36.2%, respectively. Dark peripheral dermatoscope borders and other acquisition-related artifacts were qualitatively frequent among eligible images before final selection, although artifact-specific counts were not retained. The resulting modeling cohort contained 1,199 unique patients and lesions and was near balanced: 48.2% benign and 51.8% malignant. There were no repeated patients or lesions across folds.

### 3.2 Clinical composition of the selected cohort

The malignant group was approximately 16 years older on average than the benign group (approximately 64 versus 48 years). The benign images were distributed principally across the 30–44, 45–59, and 60–75-year groups, whereas 70.5% of malignant images were in the combined 60–75 and *>* 75-year groups. All images were allocated to an age category.

Men accounted for 47.8% of the malignant group and 36.0% of the benign group. This difference was statistically significant (*χ*^2^ = 17.21, 1 degree of freedom, *p <* 0.001; Cramér’s *V* = 0.120). The odds of male sex were 1.63 times higher among malignant than benign records (95% CI 1.29–2.06). This association describes the selected cohort and should not be interpreted as an etiologic estimate.

Anatomic-site distributions differed between classes (*χ*^2^ = 117.61, 4 degrees of freedom, *p <* 0.001; Cramér’s *V* = 0.313). Palms/soles represented 58.0% of malignant images and 32.4% of benign images, corresponding to a prevalence ratio of 1.79 (95% CI 1.56–2.05).

Among malignant images, the reported subtype distribution was 48.3% melanoma in situ, 42.3% invasive melanoma, 8.2% melanoma not otherwise specified, and 1.2% metastatic melanoma. Model performance was not reported separately for these subtypes.

### 3.3 Training dynamics

Training duration differed across the five folds under the supplied AUC-based early-stopping protocol (Table 2; Figure 2a–b). Fold 1 trained for 28 epochs and reached its highest ROC-AUC of 0.977 at epoch 22. Fold 4 trained for 27 epochs and reached the highest ROC-AUC among all folds, 0.983 at epoch 21. Folds 2, 3, and 5 reached their reported maxima earlier. Fold 5 reached its reported maximum at epoch 15 and terminated at epoch 16; the available training summary did not permit independent verification of the complete early-stopping patience sequence.

**Table 2.**
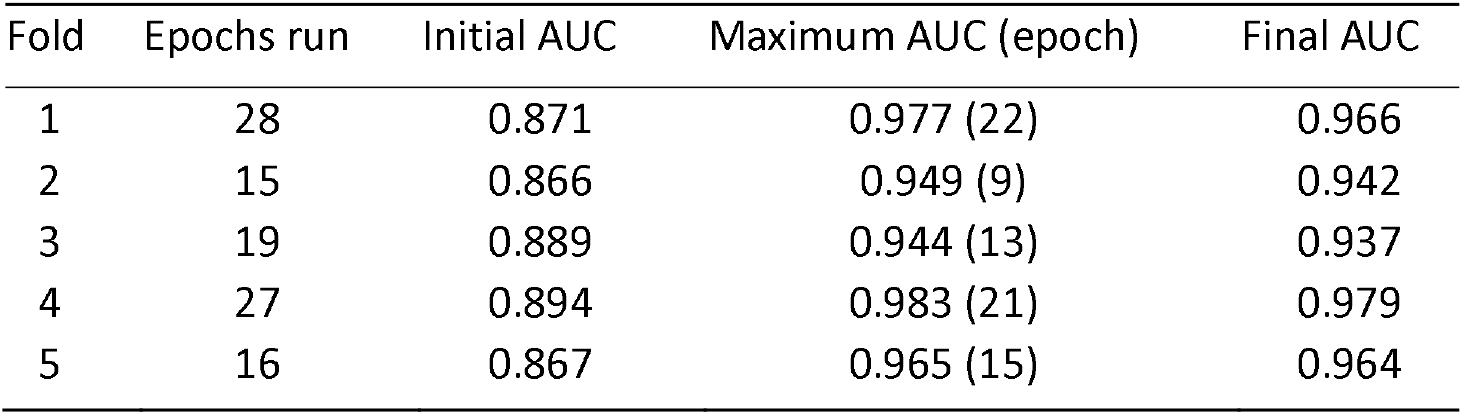
Reported fold-specific training progression. Values in the final column are those reported at the last described epoch, whereas the maximum column identifies the retained checkpoint performance.

| Fold | Epochs run | Initial AUC | Maximum AUC (epoch) | Final AUC |
| --- | --- | --- | --- | --- |
| 1 | 28 | 0.871 | 0.977 (22) | 0.966 |
| 2 | 15 | 0.866 | 0.949 (9) | 0.942 |
| 3 | 19 | 0.889 | 0.944 (13) | 0.937 |
| 4 | 27 | 0.894 | 0.983 (21) | 0.979 |
| 5 | 16 | 0.867 | 0.965 (15) | 0.964 |

For Fold 1, accuracy increased from 0.787 at epoch 1 to 0.879 at epoch 7 and 0.917 at epoch 22. ROC-AUC increased from 0.871 to 0.953 and then to 0.977 over the same epochs. At epoch 28, accuracy and ROC-AUC were 0.904 and 0.966, respectively; the earlier epoch-22 checkpoint was therefore retained. Fold 2 reached a maximum ROC-AUC of 0.949 at epoch 9 and stopped at epoch 15. Fold 3 reached 0.944 at epoch 13 and stopped at epoch 19. Fold 5 reached 0.965 at epoch 15, with a final reported ROC-AUC of 0.964 at epoch 16.

### 3.4 Cross-validation performance

Fold-specific classification results are presented in Table 3. ROC-AUC ranged from 0.9440 in Fold 3 to 0.9827 in Fold 4. Accuracy ranged from 0.845833 to 0.925000. Fold 4 had the highest accuracy, precision, recall, F1-score, and ROC-AUC among the reported fold results.

**Table 3.** Five-fold cross-validation results. Precision, recall, and F1-score are support-weighted estimates.

| Fold | Accuracy | ROC-AUC | Precision | Recall | F1-score |
| --- | --- | --- | --- | --- | --- |
| 1 | 0.916667 | 0.9767 | 0.917039 | 0.9167 | 0.916597 |
| 2 | 0.879167 | 0.9486 | 0.879993 | 0.8792 | 0.879198 |
| 3 | 0.845833 | 0.9440 | 0.862422 | 0.8458 | 0.844690 |
| 4 | 0.925000 | 0.9827 | 0.925170 | 0.9250 | 0.925021 |
| 5 | 0.899582 | 0.9654 | 0.899904 | 0.8996 | 0.899490 |

Folds 1–3 each used 959 training images (462 benign and 497 malignant) and 240 validation images (116 benign and 124 malignant). Fold 4 used 959 training images (463 benign and 496 malignant) and 240 validation images (115 benign and 125 malignant). Fold 5 used 960 training images (463 benign and 497 malignant) and 239 validation images (115 benign and 124 malignant). Exact age, sex, and anatomic-site distributions by fold were not retained; therefore, demographic distribution differences between each training and validation split could not be tabulated.

The pooled global ROC curve yielded an AUC of 0.962, and the pooled global precision–recall curve yielded an AUC of 0.969 (Figure 2c–d). The global ROC-AUC is distinct from the unweighted mean fold ROC-AUC of 0.96348 because the two values summarize performance differently.

The five fold-specific confusion matrices are shown as a single composite figure (Figure 3). Rows represent the histopathologic reference class and columns represent the predicted class. Treating malignant melanoma as positive, the cell order is TN, FP, FN, and TP from the upper-left to the lower-right corner.

**Figure 3.**
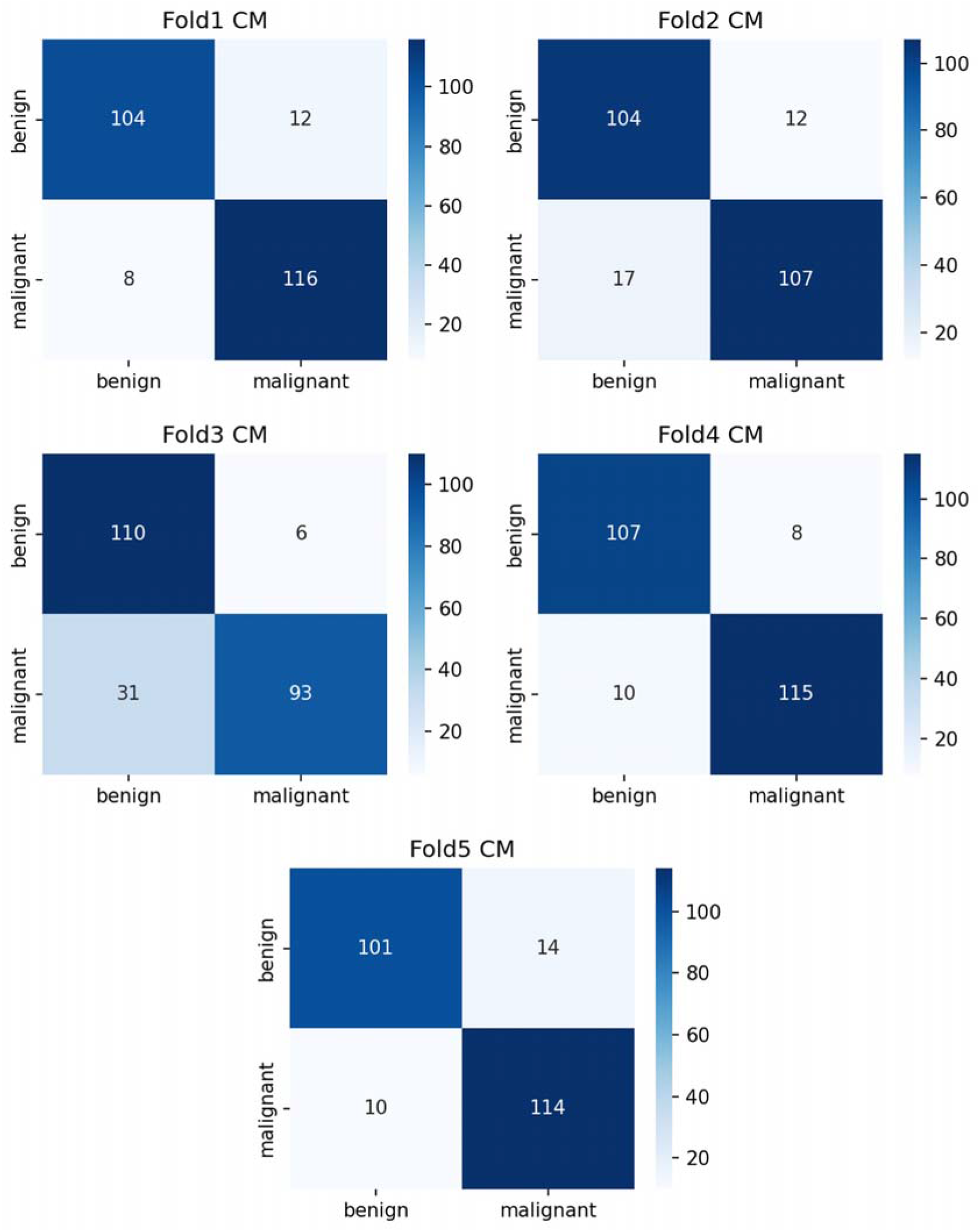
Fold-specific confusion matrices. Rows indicate the histopathologic reference class and columns indicate the predicted class. The corresponding true-negative, false-positive, false-negative, and true-positive counts were Fold 1 (104,12,8,116), Fold 2 (104,12,17,107), Fold 3 (110,6,31,93), Fold 4 (107,8,10,115), and Fold 5 (101,14,10,114).

Across folds, the recalculated mean ROC-AUC was 0.96348 ± 0.01695, and the recalculated mean accuracy was 0.89325 ± 0.03179. Mean precision, recall, and F1-score were 0.89691, 0.89326, and 0.89300, respectively (Table 4). The coefficient of variation was 1.76% for ROC-AUC and 3.56% for accuracy.

**Table 4.** Descriptive summary of fold-level performance.

| Metric | Mean | Standard deviation | Minimum | Maximum | Range |
| --- | --- | --- | --- | --- | --- |
| Accuracy | 0.89325 | 0.03179 | 0.8458 | 0.9250 | 0.0792 |
| ROC-AUC | 0.96348 | 0.01695 | 0.9440 | 0.9827 | 0.0387 |
| Precision | 0.89691 | 0.02592 | 0.8624 | 0.9252 | 0.0627 |
| Recall | 0.89326 | 0.03181 | 0.8458 | 0.9250 | 0.0792 |
| F1-score | 0.89300 | 0.03220 | 0.8447 | 0.9250 | 0.0803 |

### 3.5 Fold-level confidence intervals

Using the sample standard deviations recalculated from the fold table and a *t* critical value of 2.776, the descriptive 95% confidence interval for mean ROC-AUC was 0.9424–0.9845. The corresponding interval for mean accuracy was 0.8538–0.9327. Intervals for precision, recall, and F1-score are shown in Table 5.

**Table 5.**
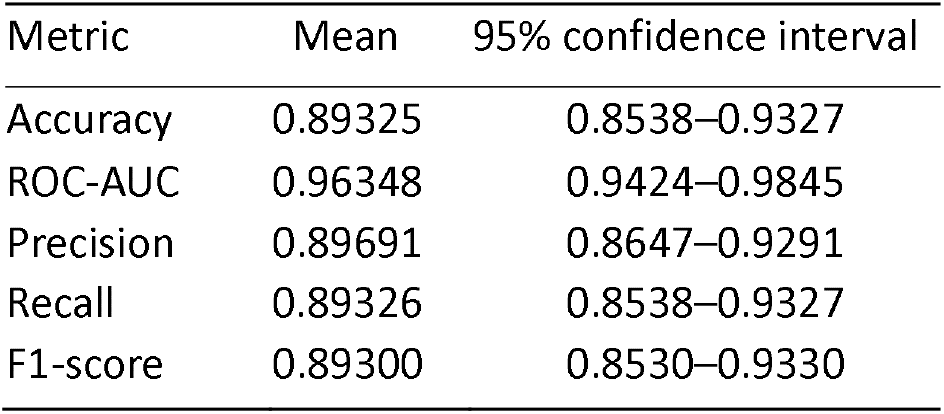
Reported fold-level mean performance and descriptive 95% confidence intervals.

These intervals summarize variability among the five reported folds. Because the training sets overlap across folds, they are presented as descriptive fold-level intervals rather than as evidence from five independent cohorts.

### 3.6 Operating threshold and interpretability results

Predicted labels were assigned by the maximum mean TTA probability; no separately optimized clinical operating threshold was reported. Summation of the five confusion matrices yielded 526 true negatives, 52 false positives, 76 false negatives, and 545 true positives. The corresponding pooled threshold-dependent estimates were 89.32% accuracy, 87.76% sensitivity, 91.00% specificity, 91.29% positive predictive value, and 87.38% negative predictive value. Across individual folds, sensitivity ranged from 75.0% to 93.5% and specificity ranged from 87.8% to 94.8%.

The matrices contained 578 benign and 621 malignant reference images in total and therefore exactly reproduced the reported cohort composition. Fold-wise accuracies and support-weighted precision, recall, and F1-scores recalculated from these counts matched Table 3 to the reported rounding precision. ROC-AUC cannot be independently reconstructed from hard-label confusion matrices because it requires the underlying continuous malignant-class probabilities; the reported ROCAUC values therefore remain based on the saved probability outputs. Global PR-AUC was 0.969 (Figure 2d); fold-specific PR-AUC and calibration results were unavailable. No subgroup-fairness, contributor-cluster, model-updating, recalibration, or decision-curve results were available.

In the two supplied false-positive Fold 1 examples, high-intensity Grad-CAM activation extended into noncutaneous dark regions at the image periphery, including the upper corners in Figure 4a and the lower-right dermatoscope rim in Figure 4b. This qualitative pattern suggests that residual dermatoscope-border artifacts may have served as spurious predictive content and may have contributed to the malignant predictions. Because only two representative errors were examined and no artifact-masking or cropping ablation was performed, the errors cannot be causally attributed to these artifacts.

**Figure 4.**
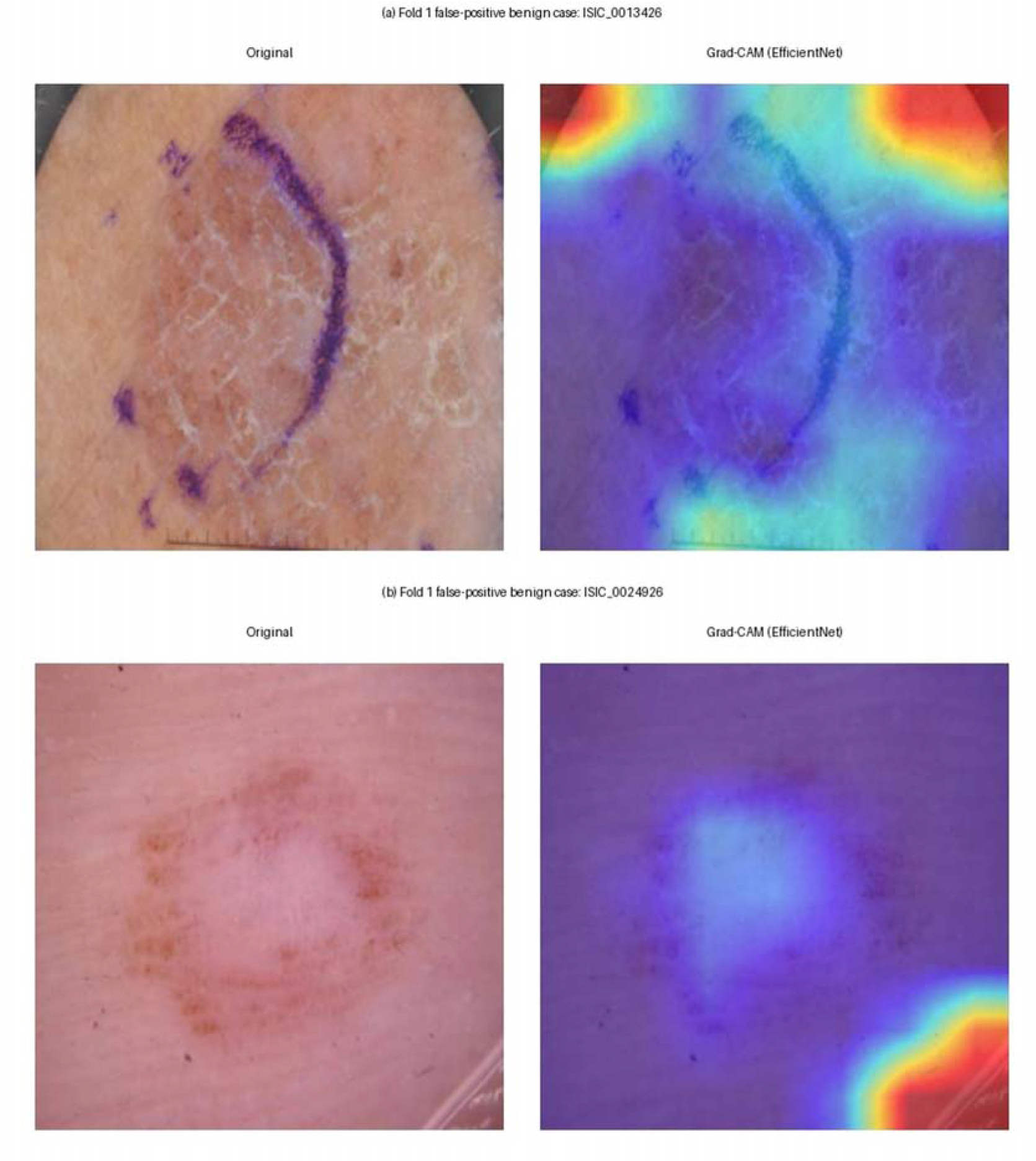
Representative Fold 1 false-positive cases. For each histopathologically benign lesion, the original dermoscopic image is shown beside its gradient-weighted class activation mapping (Grad-CAM) overlay from the EfficientNetB0 branch: (a) ISIC_0013426 and (b) ISIC_0024926. Both lesions were predicted as malignant. In (a), strong activation extends into the dark noncutaneous regions at the upper image periphery; in (b), the dominant peripheral activation includes the lower-right dermatoscope rim. These patterns suggest that residual acquisition artifacts may have acted as spurious predictive cues and may have contributed to the false-positive predictions. Grad-CAM alone cannot establish causality, and the maps were not formally scored by dermatologists.

In the two supplied correctly classified Fold 1 examples, the high-intensity Grad-CAM regions coincided predominantly with the visible lesion, its margins, and the immediate perilesional transition (Figure 5). This pattern is consistent with the EfficientNetB0 branch using lesion-boundary information and adjacent skin changes in these examples. The observation is limited to the displayed cases because systematic localization scoring and dermatologist assessment were unavailable.

**Figure 5.**
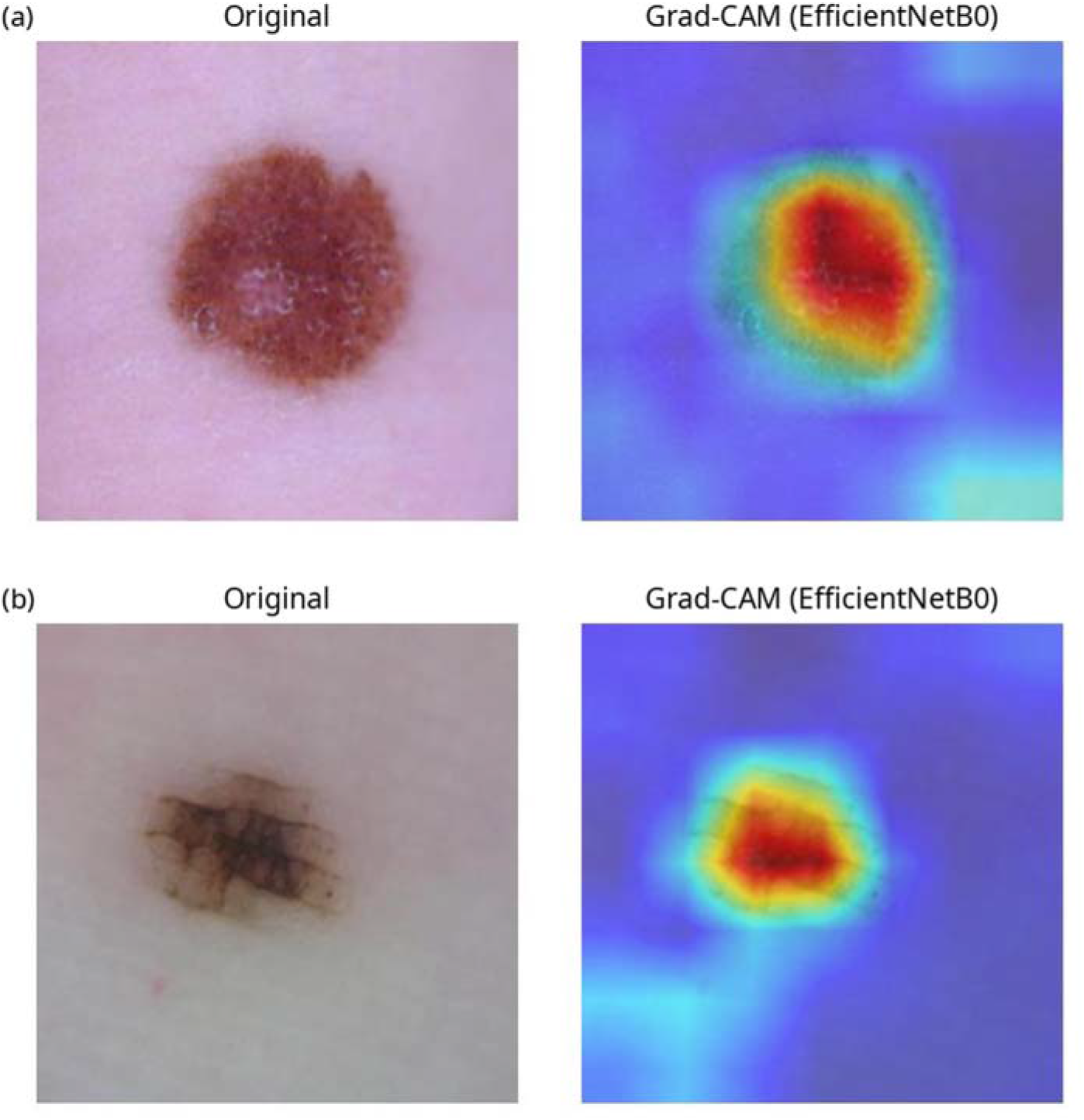
Representative Fold 1 correctly classified cases shown as vertically stacked panels. For each case, the original dermoscopic image is shown on the left and the corresponding gradient-weighted class activation mapping (Grad-CAM) overlay from the EfficientNetB0 branch on the right: (a) ISIC_0027034 and (b) ISIC_0064364. In both examples, the highest-activation red–yellow regions are concentrated predominantly over the visible lesion, its margins, and the immediate perilesional transition, whereas most distant surrounding skin shows lower activation. This distribution is consistent with the convolutional branch using lesion-boundary information and adjacent skin changes in these correctly classified cases. The overlays identify regions contributing strongly to the model output but do not establish causal clinical reasoning and were not formally scored by dermatologists.

The audit code was configured to flag every disagreement between the model prediction and the histopathologic label in the validation folds and to preserve those cases for visual review. In the two-dimensional UMAP projection, benign observations were concentrated mainly toward the left side and malignant observations mainly toward the right side, while several central and peripheral regions contained overlapping or intermixed classes (Figure 6). This pattern indicates partial class-wise organization of the fused representation rather than complete separability. Because UMAP is a nonlinear, stochastic dimensionality-reduction method, the visualization is exploratory and does not constitute an independent estimate of classification performance.

**Figure 6.**
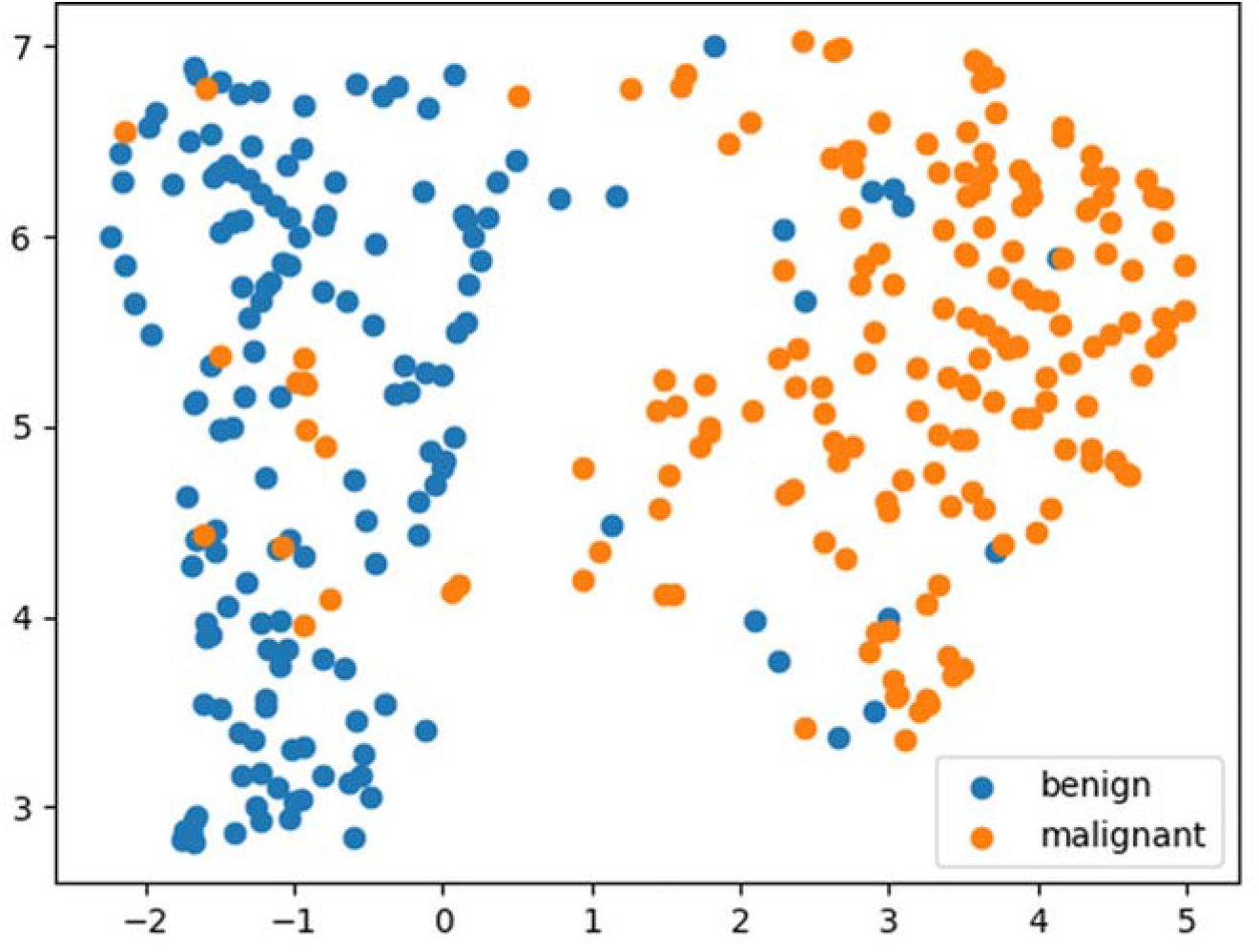
Two-dimensional Uniform Manifold Approximation and Projection (UMAP) of the fused 2,048dimensional EfficientNetB0–Swin-T representations. Each point represents an analyzed image, with histopathologically benign lesions shown in blue and malignant melanoma shown in orange. The projected representations exhibit broad class-wise organization together with overlapping transition regions. The axes are dimensionless UMAP coordinates; the plot is an exploratory representation analysis rather than a performance metric.

The generated image-level misclassification summary and complete set of audit heat maps were unavailable. Accordingly, this study does not claim that review of these cases increased accuracy.

## 4 Discussion

### 4.1 Principal findings

The proposed heterogeneous ensemble was developed using 1,199 histopathologically verified dermoscopic images selected from 552,869 screened ISIC records. The final cohort included 578 benign melanocytic lesions and 621 malignant melanoma images. Across stratified five-fold internal validation, the model achieved a recalculated mean ROC-AUC of 0.96348 and a mean accuracy of 0.89325. Global curve analysis yielded ROC-AUC 0.962 and PR-AUC 0.969 (Figure 2c–d). The pooled confusion matrices yielded sensitivity of 87.76% and specificity of 91.00%. ROC-AUC varied by 0.0387 from the lowest to the highest fold, while accuracy varied by 0.0792. The highest validation performance was obtained in Fold 4, with ROC-AUC 0.9827 and accuracy 0.9250.

Performance was not uniform across partitions. Folds 1 and 4 continued improving over longer training trajectories, whereas Folds 2, 3, and 5 reached their reported maxima earlier. The earlystopping mechanism retained the highest-AUC checkpoint rather than the final epoch, preventing the late declines observed in several folds from determining the reported fold results. For example, Fold 1 reached ROC-AUC 0.977 at epoch 22 but ended at 0.966 at epoch 28.

The model fused a 1,280-dimensional convolutional representation with a 768-dimensional shiftedwindow transformer representation. This design allowed both branches to contribute directly to the final two-class decision through a compact linear head. Selection of the B0 and Tiny variants was deliberate: the design objective was to combine local texture sensitivity and broader contextual modeling while avoiding the additional computational burden of larger backbone variants. In this sense, the architecture was intended to maximize predictive efficiency—accuracy and representational complementarity relative to computational demand and to remain practical for heterogeneous dermoscopic inputs. The reported results evaluate the complete training system, which also incorporated weighted sampling, image augmentation, mixup, label smoothing, dropout, and TTA. Because no component wise ablation, runtime benchmark, or energy measurement was supplied, the independent contribution of each backbone and any actual energy advantage cannot be quantified.

### 4.2 Context within published dermoscopic classification systems

Table 6 places the present results alongside selected published CNN, transformer, and hybrid systems. The studies used different datasets, class definitions, sample sizes, split strategies, and outcome averaging; their numerical values are therefore contextual rather than a head-to-head ranking. In particular, multiclass HAM10000 or ISIC 2018 performance cannot be directly compared with the present histopathology-filtered binary cohort.

**Table 6.** Contextual comparison with selected published dermoscopic classification systems. NR, not reported in the cited summary; values are not directly comparable across different datasets and protocols.

| Study | Model | Dataset/task | Validation | Accuracy | ROC-AUC |
| --- | --- | --- | --- | --- | --- |
| Present Study | EfficientNetB0–Swin-T | ISIC subset, binary, $n = 1,199$ | Stratified 5-fold | 89.33% | 0.9635 |
| Shen et al. [9] | Three-CNN ensemble | ISIC 2016, binary | Study-specific split | 86.91% | NR |
| Nie et al. [10] | CNN–Transformer hybrid | ISIC 2018, seven classes | Study-specific split | 89.48% | 0.96 micro; 0.90 macro |
| Sarker and Yeafi [11] | EfficientNet–Swin hybrid | HAM10000, multiclass | Study-specific split | 98% | NR |
| Lilhore et al. [12] | Explainable hybrid framework | Multiple dermoscopic datasets | Cross-dataset analyses | $98.7\% \pm 0.1\%$ | 0.995–0.998 |

The wider comparison landscape also includes output-level CNN ensembles [4], grouped deepmodel ensembles [5], CNN–vision-transformer deep ensembles [6], explainable CNN–transformer fusion systems [7], and computationally optimized EfficientNetB0 variants evaluated across several dermoscopic datasets [8]. These studies support the architectural rationale for complementary feature fusion, but their reported results do not establish superiority of the present configuration because the cohorts and validation protocols differ.

The present model’s ROC-AUC is competitive within this heterogeneous literature, but superiority cannot be inferred. A defensible novelty claim rests on the complete configuration histopathology filtered near balanced cohort, EfficientNetB0–Swin-T feature fusion, mixup, weighted sampling, label smoothing, five-view TTA, error auditing, and planned complementary explainability rather than on asserting that no related CNN–Transformer system exists. Direct single-backbone and component-removal experiments on the same folds remain necessary to establish incremental benefit.

### 4.3 Clinical interpretation and potential medical role

ROC-AUC describes ranking discrimination across thresholds, whereas clinical use requires an explicit operating threshold and the corresponding balance between missed malignant cases and false alarms. In the present task, a false negative represents a malignant image classified as benign. At the implemented maximum-probability decision rule, the pooled matrices identified 76 false-negative malignant images and 52 false-positive benign images, with sensitivity of 87.76% and specificity of 91.00%. These values characterize the retrospective operating point but do not establish an optimized clinical threshold.

The mean ROC-AUC of 0.96348 and mean accuracy of 0.89325 indicate high internal discrimination within the selected ISIC cohort. A medically appropriate use case would be as an adjunctive decision-support layer that prioritizes suspicious lesions for dermatologist review, highlights cases requiring closer inspection, and provides a reproducible second reading. Histopathology remains the reference standard in the present cohort, and the model output should not be interpreted as an autonomous diagnosis.

In any future implementation, a trained clinician would need to verify that the input is a dermoscopic RGB image, that the lesion is adequately visible and in focus, and that borders, rulers, bubbles, hair, or other artifacts do not dominate the frame. The current system has no validated automated image-quality gate or missing-input mechanism; unsuitable images should be rejected or reacquired. The clinician would remain responsible for integrating the malignant probability and any heat map with clinical context and for all referral, surveillance, biopsy, and diagnostic decisions.

Similarly, selecting Fold 4 as the global best checkpoint identifies the model state with the highest reported internal validation ROC-AUC. It does not constitute evaluation on an independent external test set. Claims regarding deployment, clinical utility, transportability, or equivalence to clinician assessment are outside the supplied evidence.

### 4.4 Generalizability and technical transferability

The model accepts standard three-channel dermoscopic images and uses two widely available pretrained backbones; therefore, the architecture is technically transferable to other dermoscopic datasets after appropriate preprocessing, retraining, and validation. The multi-contributor ISIC source exposes development to images originating from several institutions and countries. This source heterogeneity is a strength, but it is not independent external validation because contributor images were mixed across the same archive-derived folds and no previously unseen center was held out.

The internal validation design and its remaining transportability gaps are summarized in Table 7. Universal or cross-domain applicability must be tested prospectively using independent cohorts that differ by institution, geography, dermatoscope, image resolution, and clinical case mix. Domain-shift analysis, subgroup reporting, calibration assessment, and out-of-distribution detection would be required before describing the system as generally applicable.

**Table 7.**
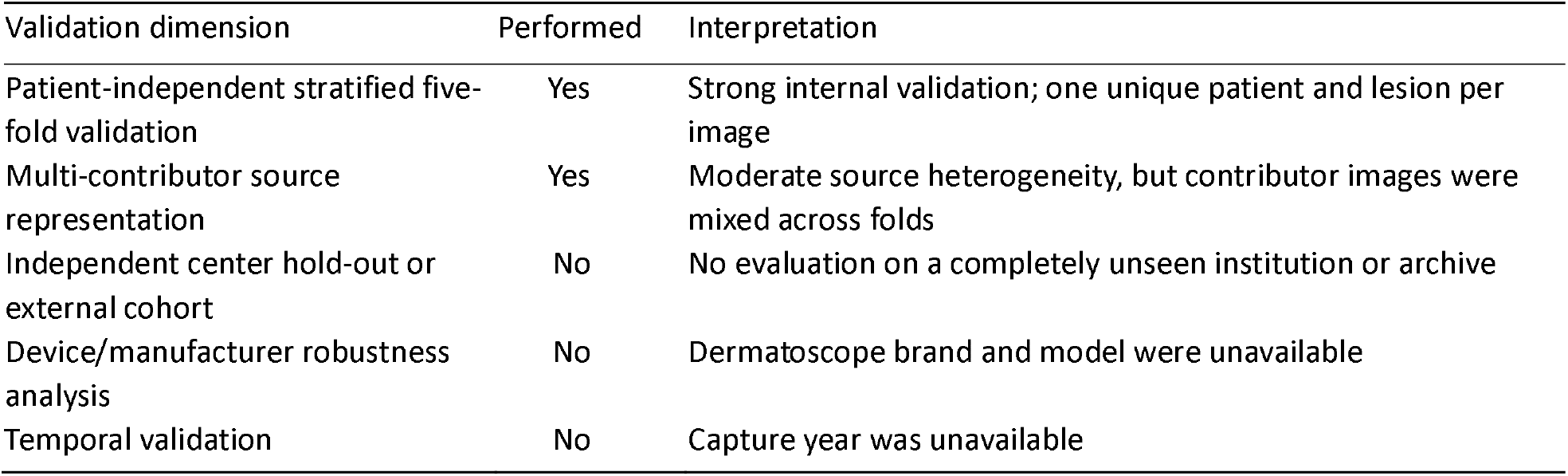
Validation scope and interpretation.

Because skin tone, race, ethnicity, socioeconomic status, contributor, and device information were unavailable, the present analysis cannot establish equitable performance. Future external validation should prespecify subgroup sample sizes and report discrimination, calibration, error rates, and potential clinical consequences across relevant populations.

### 4.5 Explainability and systematic error reduction

The proposed workflow combines two complementary interpretability levels. Grad-CAM localizes image regions contributing to a prediction, whereas UMAP shows the organization of fused representations. In the representative false-positive outputs, activation extended into dark noncutaneous dermatoscope-border regions (Figure 4); in the correctly classified outputs, activation was concentrated predominantly on the lesions, their margins, and adjacent perilesional transitions (Figure 5). This contrast supports the hypothesis that peripheral acquisition artifacts can provide spurious cues, while lesion-centered localization may accompany correct classification, but neither relationship was quantified or tested causally. The supplied UMAP projection demonstrated broad separation of benign and malignant observations, with residual overlap and isolated points across the projected space (Figure 6). These overlapping neighborhoods may contain visually ambiguous or atypical lesions, but image-level linkage and expert review would be required to establish their clinical meaning. Together with the correct/incorrect prediction reports, these tools create an auditable framework in which difficult lesions, image artifacts, atypical morphology, and recurrent failure patterns can be reviewed.

Because the validation labels were known, all prediction–reference disagreements could be flagged and collected. These cases could subsequently support dermatologist review, hard-example mining, targeted data augmentation, threshold adjustment, or retraining. Such a feedback loop has the potential to improve later model versions, but improvement was not tested in the supplied experiment. In prospective unlabeled data, the algorithm cannot know that a specific prediction is erroneous without an independent reference or human adjudication.

### 4.6 Image-processing refinements and future development

The current preprocessing pipeline was deliberately limited to resizing, geometric and color augmentation, coarse dropout, and fixed channel normalization (Table 8). The instrument only archive field describes image provenance/manipulation and should not be interpreted as explicit computational removal of hair, rulers, dark dermatoscope borders, or other artifacts. Robustness to varying illumination and artifacts was encouraged through stochastic augmentation, but performance was not measured separately across such conditions. The qualitative false-positive Grad-CAM findings indicate that residual peripheral artifacts warrant explicit evaluation rather than reliance on augmentation alone.

**Table 8.** Image-processing components implemented or reserved for future evaluation.

| Processing component | Used | Current implementation and interpretation |
| --- | --- | --- |
| Resize and intensity normalization | Yes | 224 x 224 single scale input; channel mean and SD of 0.5 |
| Geometric and photometric augmentation | Yes | Flip, right-angle rotation, and color jitter for training-time robustness |
| Occlusion augmentation | Yes | Coarse dropout to simulate local masking |
| Lesion segmentation or centered cropping | No | Candidate ablation for reducing background influence |
| Explicit hair/ruler/airbubble/dermatoscope-border removal | No | Candidate artifact-specific preprocessing |
| CLAHE, color constancy, or illumination correction | No | No correction beyond fixed normalization |
| Device-specific harmonization | No | Device metadata were unavailable; requires device-labeled validation |
| Higher-resolution or multi-scale input | No | Candidate approach for preserving fine dermoscopic structures |

Further development could therefore evaluate lesion segmentation or lesion-centered cropping, masking of dark dermatoscope rims and vignettes, hair and ruler-mark suppression, illumination and color-constancy correction, air-bubble handling, device-specific color harmonization, multi-scale inputs, and higher-resolution fine-tuning. More systematic image-quality review and artifact-aware preprocessing could reduce spurious peripheral activation and produce more stable, lesion-focused predictions; whether these changes improve accuracy must be established through prespecified ablation experiments rather than assumed from the displayed examples.

Any preprocessing extension should be assessed through prespecified ablation experiments on the same patient-separated folds. Future optimization should also examine sensitivity-oriented threshold selection, probability calibration, uncertainty estimation, and training strategies that place greater emphasis on histopathologically confirmed false-negative melanoma cases.

The intended roles of the complete system’s components are summarized in Table 9. No componentremoval experiment was performed; accordingly, numerical improvement estimates for individual components are not reported as study findings.

**Table 9.** Intended component roles and empirical ablation status.

| Component | Intended role | Same-cohort ablation evidence |
| --- | --- | --- |
| EfficientNetB0 | Local convolutional morphology and texture representation | Not tested separately |
| Swin-T | Hierarchical shifted-window contextual representation | Not tested separately |
| Feature concatenation | Joint use of complementary backbone representations | No alternative fusion tested |
| Mixup | Smoother decision boundary and regularization | No no-mixup comparison |
| Five-view TTA | Averaging across augmented inference views | No single-view comparison |
| Label smoothing | Reduction of overconfident targets | No unsmoothed comparison |
| Weighted sampling | Balanced exposure to the two classes | No unweighted comparison |

### 4.7 Strengths

The study used an explicitly filtered, histopathologically verified dermoscopic cohort from a publicly accessible, multi-contributor archive. A dermatologist performed image-quality and near-duplicate review, and the final cohort contained exactly one image for each of 1,199 patients and lesions. The benign and malignant groups were near balanced. The modeling design also incorporated patient-independent stratified five-fold validation, independent training within each fold, inversefrequency sampling, a heterogeneous feature-fusion architecture, combined input- and target-level regularization, AUC-based checkpoint selection, and probability averaging over five augmented views. Reporting fold-specific values, rather than only the best fold, made cross-partition variability visible.

The analysis included image-level and representation-level interpretability using Grad-CAM and UMAP. Representative Grad-CAM false-positive and correctly classified examples documented where the convolutional branch focused (Figures 4 and 5), while the UMAP visualization documented the low-dimensional arrangement of the fused representations (Figure 6).

### 4.8 Limitations

Although the final image count, class distribution, eight mandatory fields, dermatologist quality review, and one-patient/one-lesion design were specified, several cohort details remain unresolved. The archive extraction date, contributor-specific counts, exact per-image ISIC identifiers and licenses, reason-specific quality-exclusion counts, capture dates, and acquisition-device metadata were unavailable. The substantial difference in selection rates between the benign and malignant branches may introduce selection bias.

The available identifier and manual-review process excluded within-cohort duplicates and repeated patients, but potential overlap between the selected ISIC images and data used during ImageNet pretraining or other prior model development was not assessed.

Third, the held-out fold was used both for early stopping and checkpoint selection and for estimating fold performance; without a nested inner validation set or an independent test set, the reported internal performance may be optimistic. Fourth, there was no independent external or temporally separated test cohort. Device brands were unavailable, and no device-robustness or geographic subgroup analysis was reported. Fifth, although confusion matrices and threshold-specific sensitivity and specificity were available, the decision threshold was not clinically optimized or externally validated. Sixth, global PR-AUC was available, but fold-specific PR-AUC, calibration, confidence intervals based on image-level predictions, and statistical comparisons with baseline or single-backbone models were unavailable. Seventh, representative Grad-CAM and UMAP visualizations were available, but a systematic expert assessment of the activation maps, artifact-stratified performance, artifact-masking ablation, the exact UMAP hyperparameters and random seed, imagelevel UMAP coordinates, and quantitative representation analyses were unavailable. The apparent peripheral-artifact activation in two false-positive examples is therefore hypothesis-generating and should not be generalized to all model errors. Finally, the local workstation’s exact operating system and hardware specifications, floating-point operation count, inference latency, electrical energy consumption, carbon emissions, total training time, and a versioned code archive remain necessary for full reproducibility. Consequently, computational and energy efficiency should be interpreted as architecture-selection objectives, not validated performance claims.

The multi-contributor origin of the images does not by itself establish external validity because all images were filtered from a single archive and evaluated only by internal cross-validation. The study therefore supports high internal performance and technical portability, not universal clinical generalizability.

## 5 Conclusions

An ImageNet-pretrained EfficientNetB0–Swin-T feature-fusion ensemble was developed using 1,199 histopathologically verified dermoscopic images representing 1,199 unique patients and lesions from the ISIC Archive. In patient-independent stratified five-fold internal validation, the system achieved recalculated mean ROC-AUC 0.96348±0.01695 and mean accuracy 0.89325±0.03179. The integrated classification, explainability, and error-audit workflow provides a technically transferable platform for subsequent dermatologic decision-support research. Its accuracy may be further improved through systematic review of labeled errors, refined image preprocessing, calibration, and sensitivity-oriented optimization. These possibilities remain hypotheses until evaluated experimentally; population- and device-diverse independent external validation is required before universal or clinical-use claims can be supported.

## Supporting information

TRIPOD+AI and CLAIM Checklists

## Ethics Statement

The broader study protocol, titled “Dermoscopy-Based Melanoma Detection: Artificial Intelligence, Dermatologist Performance, and the Effect of Joint Decision-Making” (original title: “Dermoskopide Melanom Tespiti: Yapay Zekâ, Dermatolog Performansı ve Birlikte Karar Vermenin Etkisi”), was reviewed by the Mardin Artuklu University Non-Interventional Clinical Research Ethics Committee. Following application no. 250649 dated April 3, 2026, the committee approved the study unanimously at session no. 4 on April 7, 2026 (Decision no. 2026/4-15). Mehmet Tarık Baran was named as the researcher. This secondary analysis used publicly accessible, de-identified dermoscopic images and involved no direct participant recruitment or intervention.

## Informed Consent Statement

Individual informed consent was not applicable because this secondary analysis used publicly accessible, de-identified dermoscopic images and involved no direct participant recruitment or intervention.

## Consent for Publication

Not applicable. This manuscript contains no identifiable personal information or identifiable participant images.

## Data Availability Statement

The source images and metadata are publicly available through the International Skin Imaging Collaboration Archive (https://www.isic-archive.com/). Individual images remain subject to the license and attribution requirements attached to each ISIC record. The filtered 1,199-image cohort manifest, applied query criteria, and associated metadata supporting the findings of this study are available from the corresponding author upon reasonable request and subject to the applicable ISIC terms of use.

## Code Availability Statement

The code used for data preprocessing, model training, cross-validation, performance evaluation, and interpretability analyses, together with the model definition, preprocessing configuration, fold assignments, and fold-specific checkpoints needed to reproduce the reported predictions, is available from the corresponding author upon reasonable request. No versioned public repository or deployment interface is currently available. Reuse remains subject to the licenses attached to the source images and pretrained weights.

## Protocol Availability Statement

No separate prospective prediction-model protocol was prepared. The broader ethics-approved study protocol described above is available from the corresponding author upon reasonable request.

## Study Registration

This retrospective prediction-model study was not registered.

## Patient and Public Involvement Statement

Patients and members of the public were not involved in the design, conduct, reporting, interpretation, or dissemination planning of this secondary analysis.

## Author Contributions

Mehmet Tarık Baran: Conceptualization; methodology; data curation; software; validation; formal analysis; visualization; writing–original draft; writing–review and editing. Ömer Karakoyun: Validation; writing–review and editing. All authors read and approved the final manuscript.

## Acknowledgments

The authors gratefully acknowledge the International Skin Imaging Collaboration and the contributing institutions and investigators for making the dermoscopic images and associated metadata available for research. Use of the source material is subject to the license and attribution requirements of the corresponding ISIC records.

## Generative Artificial Intelligence Disclosure

During manuscript preparation, a generative artificial intelligence assistant was used in a limited capacity for language refinement and for information-seeking support related to manuscript verification and code-debugging concepts. The tool was not used to generate or alter study images, reference labels, model training data, model outputs, statistical results, or clinical decisions. The authors reviewed and verified the manuscript content and assume full responsibility for its accuracy, integrity, and final wording.

## References

[1] Arnold M, Singh D, Laversanne M, et al. Global burden of cutaneous melanoma in 2020 and projections to 2040. JAMA Dermatol. 2022;158(5):495–503. doi:10.1001/jamadermatol.2022.0160.

[2] Tan M, Le QV. EfficientNet: rethinking model scaling for convolutional neural networks. In: Proceedings of Machine Learning Research. 2019;97:6105–6114.

[3] Liu Z, Lin Y, Cao Y, et al. Swin Transformer: hierarchical vision transformer using shifted windows. In: Proceedings of the IEEE/CVF International Conference on Computer Vision; 2021:10012–10022. doi:10.1109/ICCV48922.2021.00986.

[4] Harangi B. Skin lesion classification with ensembles of deep convolutional neural networks. J Biomed Inform. 2018;86:25–32. doi:10.1016/j.jbi.2018.08.006.

[5] Guergueb T, Akhloufi MA. Skin cancer detection using ensemble learning and grouping of deep models. In: Proceedings of the 19th International Conference on Content-Based Multimedia Indexing (CBMI); 2022:121–125. doi:10.1145/3549555.3549584.

[6] Chiu TM, Li YC, Chi IC, Tseng MH. Deep ensemble learning with convolution neural networks and vision transformers for skin lesion classification. SSRN [Preprint]. Posted October 26, 2024. doi:10.2139/ssrn.4984829.

[7] Ali A, Shahbaz H, Damaševičius R. xCViT: improved vision transformer network with fusion of CNN and Xception for skin disease recognition with explainable AI. Comput Mater Contin. 2025;83(1):1367–1398. doi:10.32604/cmc.2025.059301.

[8] Altaf AR, Altaf A, Rehman FU. Modified EfficientNet-B0 architecture optimized with quantumbehaved algorithm for skin cancer lesion assessment. Diagnostics (Basel). 2025;15(24):3245. doi:10.3390/diagnostics15243245.

[9] Shen X, Wei L, Tang S. Dermoscopic image classification method using an ensemble of fine-tuned convolutional neural networks. Sensors (Basel). 2022;22(11):4147. doi:10.3390/s22114147.

[10] Nie Y, Sommella P, Carratù M, O’Nils M, Lundgren J. A deep CNN Transformer hybrid model for skin lesion classification of dermoscopic images using focal loss. Diagnostics (Basel). 2023;13(1):72. doi:10.3390/diagnostics13010072.

[11] Sarker L, Yeafi A. EF-SwinNet: a hybrid EfficientNet–Swin Transformer model for skin cancer classification. In: 2024 International Conference on Recent Progresses in Science, Engineering and Technology (ICRPSET). IEEE; 2024:1–4. doi:10.1109/ICRPSET64863.2024.10955919.

[12] Lilhore UK, Anitha D, Priya R, et al. Adaptive hybrid AI framework for robust and explainable skin lesion segmentation and melanoma detection. Int J Comput Intell Syst. 2026;19:112. doi:10.1007/s44196-026-01233-y.

[13] Adamson AS, Smith A. Machine learning and health care disparities in dermatology. JAMA Dermatol. 2018;154(11):1247–1248. doi:10.1001/jamadermatol.2018.2348.

[14] Daneshjou R, Vodrahalli K, Novoa RA, et al. Disparities in dermatology AI performance on a diverse, curated clinical image set. Sci Adv. 2022;8(32):eabq6147. doi:10.1126/sciadv.abq6147.

[15] Tejani AS, Klontzas ME, Gatti AA, et al. Checklist for Artificial Intelligence in Medical Imaging (CLAIM): 2024 update. Radiol Artif Intell. 2024;6(4):e240300. doi:10.1148/ryai.240300.

[16] Collins GS, Moons KGM, Dhiman P, et al. TRIPOD+AI statement: updated guidance for reporting clinical prediction models that use regression or machine learning methods. BMJ. 2024;385:e078378. doi:10.1136/bmj-2023-078378.

[17] Selvaraju RR, Cogswell M, Das A, et al. Grad-CAM: visual explanations from deep networks via gradient-based localization. In: Proceedings of the IEEE International Conference on Computer Vision; 2017:618–626. doi:10.1109/ICCV.2017.74.

[18] McInnes L, Healy J, Saul N, Großberger L. UMAP: Uniform Manifold Approximation and Projection. J Open Source Softw. 2018;3(29):861. doi:10.21105/joss.00861.

