## Supplementary material for "Skin Cancer Classification Using Explainable Artificial Intelligence With an Ensemble Model and Rigorous Leakage Free Validation": TRIPOD+AI and CLAIM Checklists

**TRIPOD+AI Reporting Checklist**

**Prepared for submission to medRxiv**

### Assessment basis

This completed checklist maps every TRIPOD+AI subitem to evidence in the uploaded 34-page manuscript. Page references use the last saved manuscript pagination. Status reflects reporting completeness, not methodological quality. Statements that an analysis was not performed or information was unavailable remain explicit.

**Summary:** 31 fully reported; 16 partially reported; 5 not applicable/not performed (52 subitems).

**Legend:** ✓ Fully reported	◀ Partially reported	N/A – not performed/applicable.

### Main TRIPOD+AI checklist

| **Section/topic** | **Item** | **TRIPOD+AI requirement Status** | | **Manuscript evidence / assessment** | **Page(s)** |
| --- | --- | --- | --- | --- | --- |
| **TITLE** | **1** | Identify model study, target population, and outcome. | ◀ **Partially reported** | Title identifies AI skin-cancer classification, ensemble model, and leakage-free validation; exact dermoscopic target population and binary outcome are completed in the Abstract/Introduction. | **1–2** |
| **ABSTRACT** | **2** | Report according to TRIPOD+AI for Abstracts. | ◀ **Partially reported** | Structured abstract reports data, sample, predictors, reference, model, internal validation, results, limitations, intended role, and non-registration. Calibration is stated unavailable. | **1** |
| **INTRODUCTION –**  **Background** | **3a** | Explain healthcare context, rationale, and existing models. | ✓ **Fully reported** | Melanoma burden, clinical consequences of errors, need for calibration/interpretability, and prior CNN, transformer, ensemble, and XAI work are described. | **2–3** |
| **INTRODUCTION –**  **Background** | **3b** | Describe target population, intended purpose, pathway, and users. | ✓ **Fully reported** | Specialist/referral dermoscopy patients; dermatologists/trained clinicians; second-reading support for prioritization, referral, surveillance, or biopsy; not autonomous diagnosis. | **2** |
| **INTRODUCTION –**  **Background** | **3c** | Describe known health inequalities. | ✓ **Fully reported** | Potential inequalities across skin tone, demographics, subtype, devices, and settings are discussed; unavailable fairness metadata are disclosed. | **3, 26, 29** |
| **INTRODUCTION –**  **Objectives** | **4** | Specify model development and/or evaluation objectives. | ✓ **Fully reported** | Objective explicitly states development and internal validation of the EfficientNetB0–SwinT ensemble. | **2, 4** |
| **METHODS – Data** | **5a** | Describe data sources, rationale, and representativeness. | ✓ **Fully reported** | Retrospectively filtered public ISIC Archive supplied development/internal-validation folds; rationale and non-consecutive representativeness limits are stated. | **4, 6** |
| **METHODS – Data** | **5b** | Specify data accrual dates and follow-up. | ◀ **Partially reported** | Acquisition/accrual dates and capture year are explicitly unavailable; follow-up is not applicable to this diagnostic image study. | **4** |
| **METHODS – Partici-**  **pants** | **6a** | Describe setting, centres, and locations. | ◀ **Partially reported** | Specialist/referral context and multi-contributor ISIC source are described; centre number/location and contributor-specific counts were unavailable. | **2, 4, 6** |
| **METHODS – Partici-**  **pants** | **6b** | Describe eligibility criteria. | ✓ **Fully reported** | Dermoscopic type, histopathology, melanocytic status, diagnosis, instrument-only manipulation, metadata completeness, quality, uniqueness, and one-image-per-patient/lesion rules are specified. | **5–7** |
| **METHODS – Partici-**  **pants** | **6c** | Describe treatments and handling, if relevant. | **N/A – not per-**  **formed/applicable** | No treatment allocation or follow-up; treatment information is explicitly not applicable. | **4, 7** |
| **METHODS – Data**  **preparation** | **7** | Describe preprocessing and quality checks, including group comparability. | ◀ **Partially reported** | Quality/artifact/focus/duplicate review and complete image preprocessing are described; sociodemographic comparability could not be assessed. | **5, 8** |
| **METHODS – Out-**  **come** | **8a** | Define outcome, horizon, assessment, rationale, and group consistency. | ✓ **Fully reported** | Histopathologically verified malignant melanoma versus benign melanocytic lesion; no prognostic horizon; inability to assess group consistency is stated. | **1, 7** |
| **METHODS – Out-**  **come** | **8b** | Describe subjective-outcome assessor qualifications/demographics. | ◀ **Partially reported** | Histopathology is the reference; original pathology procedures and assessor qualifications/demographics were unavailable. | **7** |
| **METHODS – Out-**  **come** | **8c** | Report outcome-assessment blinding. | ◀ **Partially reported** | Original pathology blinding procedures were unavailable and this is explicitly reported. | **7** |
| **METHODS – Predic-**  **tors** | **9a** | Describe initial predictor choice and preselection. | ✓ **Fully reported** | Sole predictor is the RGB dermoscopic image; metadata are excluded; complementary CNN/local and transformer/contextual rationale is given. | **7, 9** |
| **METHODS – Predic-**  **tors** | **9b** | Define predictors, measurement, transformations, and blinding. | ✓ **Fully reported** | RGB conversion, dimensions, resizing, normalization, learned feature dimensions, transformations, and absence of metadata inputs are specified. | **7–9** |
| **METHODS – Predic-**  **tors** | **9c** | Describe subjective-predictor assessor qualifications/demographics. | ◀ **Partially reported** | Dermatologist quality/uniqueness review is reported; experience, demographics, and reference-label blinding were not recorded. | **7** |
| **METHODS – Sample**  **size** | **10** | Explain and justify study size. | ◀ **Partially reported** | Size arose from eligibility, completeness, quality, uniqueness, and one-patient/one-lesion rule; no formal size/precision calculation was performed. | **7** |
| **METHODS – Missing**  **data** | **11** | Describe missing-data handling and omissions. | ✓ **Fully reported** | Mandatory fields were complete; no imputation; manual-review exclusions were omitted; missing reason-specific counts are disclosed. | **5, 7** |
| **METHODS – Analysis** | **12a** | Describe data use and partitioning. | ✓ **Fully reported** | Patient/lesion-independent stratified five-fold validation, fold counts, non-overlap, trainingonly sampling, and validation-fold checkpoint use are reported. | **6, 10–12** |

Continued on next page

| **Section/topic** | **Item** | **TRIPOD+AI requirement Status Manuscript evidence / assessment** | | | **Page(s)** |
| --- | --- | --- | --- | --- | --- |
| **METHODS – Analysis** | **12b** | Describe predictor handling and standardisation. | ✓ **Fully reported** | RGB conversion, resizing, normalization, augmentation, coarse dropout, mixup, and TTA are fully specified. | **8, 11–12** |
| **METHODS – Analysis** | **12c** | Specify model, rationale, building, hyperparameters, and internal validation. | ✓ **Fully reported** | Backbones, fusion, head, initialization, optimizer, seeds, determinism, epochs, early stopping, checkpointing, TTA, and folds are detailed. | **7, 9–14** |
| **METHODS – Analysis** | **12d** | Describe cluster heterogeneity handling. | ◀ **Partially reported** | Multi-contributor heterogeneity is acknowledged; contributor counts were unavailable, contributors mixed across folds, and clustering was not modelled. | **4, 13, 26** |
| **METHODS – Analysis** | **12e** | Specify discrimination, calibration, and clinical-utility measures. | ◀ **Partially reported** | Classification/discrimination measures and plots are defined. Calibration is partially addressed in reporting but not measured; decision-curve analysis was not performed. | **12–13, 17–20** |
| **METHODS – Analysis** | **12f** | Describe updating or recalibration. | **N/A – not per-**  **formed/applicable** | No updating or probability recalibration followed validation; both are identified as future work. | **13, 29** |
| **METHODS – Analysis** | **12g** | Describe calculation of evaluation predictions. | ✓ **Fully reported** | For each image, five stochastic TTA views are evaluated. Softmax probabilities are averaged; the positive class and 0.5/maximum-probability rule are specified. | **12** |
| **METHODS – Class**  **imbalance** | **13** | Explain imbalance methods and recalibration. | ✓ **Fully reported** | Inverse-frequency weighted sampling is described; cohort is near balanced; no post-hoc recalibration and no fairness claim are stated. | **10–12** |
| **METHODS – Fairness** | **14** | Describe fairness approaches and rationale. | ◀ **Partially reported** | No demographic fairness analysis/intervention was possible; missing skin tone, race, ethnicity, socioeconomic, device, and contributor data are disclosed. | **3, 13,**  **26, 29** |
| **METHODS – Model**  **output** | **15** | Specify outputs, classifications, thresholds, and rationale. | ✓ **Fully reported** | Two-class softmax probabilities, class mapping, malignant positive class, and 0.5 rule are reported; threshold was not clinically optimized. | **9, 12** |
| **METHODS – Train-**  **ing/evaluation** | **16** | Identify development/evaluation data differences. | ✓ **Fully reported** | Same archive cohort and definitions supplied separated folds; lack of independent external/temporal data is explicit. | **4, 10,**  **26, 29** |
| **METHODS – Ethics** | **17** | Name ethics body and report consent/waiver. | ✓ **Fully reported** | Committee, application/date/session/decision are reported; consent was not applicable for public de-identified images. | **30** |
| **OPEN SCIENCE** | **18a** | State funding source and funder role. | ✓ **Fully reported** | No funding was received; consequently there was no funder role. | **31** |
| **OPEN SCIENCE** | **18b** | Declare conflicts and financial disclosures. | ✓ **Fully reported** | No relevant financial or non-financial interests are declared. | **31** |
| **OPEN SCIENCE** | **18c** | State protocol availability. | ✓ **Fully reported** | No separate prediction-model protocol; broader ethics-approved protocol is available from the corresponding author on reasonable request. | **31** |
| **OPEN SCIENCE** | **18d** | Provide registration or state non-registration. | ✓ **Fully reported** | Study is explicitly stated not registered; the Abstract also reports non-registration. | **1, 31** |
| **OPEN SCIENCE** | **18e** | Describe data availability. | ✓ **Fully reported** | ISIC images/metadata are public under licences; filtered manifest, query criteria, and metadata are available on reasonable request. | **31** |
| **OPEN SCIENCE** | **18f** | Describe code availability. | ✓ **Fully reported** | Preprocessing, training, validation, evaluation, XAI code, configuration, fold assignments, and checkpoints are available on reasonable request; no public repository. | **31** |
| **PATIENT/PUBLIC**  **INVOLVEMENT** | **19** | Describe involvement or state none. | ✓ **Fully reported** | No patient/public involvement in design, conduct, reporting, interpretation, or dissemination planning. | **31** |
| **RESULTS – Participants** | **20a** | Report participant flow and outcome counts. | ✓ **Fully reported** | Flow diagram reports 552,869 screened records through eligibility/review to 578 benign and 621 malignant unique patient-lesion images. | **5, 15** |
| **RESULTS – Participants** | **20b** | Report characteristics, dates, predictors, events, missingness, and group differences. | ◀ **Partially reported** | Age, sex, site, counts, class comparisons, and zero mandatory-field missingness are reported; dates, centres/devices, skin tone/race/ethnicity are unavailable. | **6, 15–16** |
| **RESULTS – Participants** | **20c** | Compare external-evaluation and development distributions. | **N/A – not per-**  **formed/applicable** | No external evaluation dataset exists; all folds came from the same cohort. | **4, 10,**  **26, 29** |
| **RESULTS – Model**  **development** | **21** | Report analysis sample and outcome-event numbers. | ✓ **Fully reported** | Total/class counts and fold-specific training/validation benign/malignant counts are reported; each image is one patient and lesion. | **10, 15, 17** |
| **RESULTS – Model**  **specification** | **22** | Provide the full model for independent prediction/evaluation. | ◀ **Partially reported** | Architecture, inputs, preprocessing, hyperparameters, output rule, seeds, and libraries are detailed; code/checkpoints are request-only, no public deployment object/final all-data retraining. | **7, 9–12, 31** |
| **RESULTS – Performance** | **23a** | Report performance with confidence intervals and subgroups. | ◀ **Partially reported** | Fold/pooled metrics, curves, matrices, ranges, and fold-level CIs are reported. Calibration, image-level CIs, and demographic/device/subtype subgroup results are unavailable. | **17–20** |

Continued on next page

| **Section/topic** | **Item** | **TRIPOD+AI requirement Status Manuscript evidence / assessment** | | | **Page(s)** |
| --- | --- | --- | --- | --- | --- |
| **RESULTS – Performance** | **23b** | Report cluster performance heterogeneity. | **N/A – not per-**  **formed/applicable** | Cluster heterogeneity was not examined because contributor/centre/device information was unavailable. | **13, 26** |
| **RESULTS – Updating** | **24** | Report updated model and updated performance. | **N/A – not per-**  **formed/applicable** | No updating or recalibration was performed; no updated model result exists. | **13, 29** |
| **DISCUSSION – Inter-**  **pretation** | **25** | Interpret results and fairness against objectives/prior work. | ◀ **Partially reported** | Internal performance, errors, clinical adjunct role, prior systems, and XAI are interpreted; fairness implications are discussed but not empirically evaluated. | **24–29** |
| **DISCUSSION – Limi-**  **tations** | **26** | Discuss bias, uncertainty, and generalizability limitations. | ✓ **Fully reported** | Selection bias, checkpoint reuse, absent external/device validation, calibration/subgroup gaps, limited XAI scoring, and reproducibility limits are discussed. | **28–29** |
| **DISCUSSION – Us-**  **ability** | **27a** | Describe handling of poor-quality/unavailable inputs. | ✓ **Fully reported** | Clinicians must check image type, visibility, focus, and artifacts; unsuitable images should be rejected/reacquired; no automated quality gate exists. | **25, 27** |
| **DISCUSSION – Us-**  **ability** | **27b** | State required user interaction and expertise. | ✓ **Fully reported** | Dermatologists/trained clinicians integrate probability/heat maps with context and retain responsibility for clinical decisions. | **2, 25** |
| **DISCUSSION – Us-**  **ability** | **27c** | Discuss next research for applicability/generalizability. | ✓ **Fully reported** | Calls for diverse external validation, calibration, fairness/subgroups, threshold optimization, uncertainty/OOD, artifact preprocessing, ablations, and prospective evaluation. | **26–29** |

### TRIPOD+AI for Abstracts checklist

All evidence in this table appears on manuscript page 1.

| **Section/topic** | **Item** | **TRIPOD+AI requirement** | **Status** | **Manuscript evidence / assessment** | **Page(s)** |
| --- | --- | --- | --- | --- | --- |
| **Title** | **1** | Identify study scope, population, and outcome. | ◀ **Partially reported** | Title identifies AI classification/validation; exact dermoscopic population and binary outcome appear in abstract text. | **1** |
| **Background** | **2** | Give context and rationale. | ✓ **Fully reported** | Reliable melanoma classification and leakage-aware validation rationale stated. | **1** |
| **Objectives** | **3** | Specify development/evaluation objective. | ✓ **Fully reported** | Development and internal validation are explicit. | **1** |
| **Methods** | **4** | Describe data source. | ✓ **Fully reported** | Retrospectively filtered ISIC Archive. | **1** |
| **Methods** | **5** | Describe eligibility and setting. | ◀ **Partially reported** | Histopathology, unique images, screening, filtering, review, and counts summarized; centre details unavailable. | **1** |
| **Methods** | **6** | Specify outcome and horizon. | ✓ **Fully reported** | Benign melanocytic lesion versus malignant melanoma; no prognostic horizon. | **1** |
| **Methods** | **7** | Specify model and validation. | ✓ **Fully reported** | EfficientNetB0–Swin-T fusion, training methods, and patient-independent five-fold validation summarized. | **1** |
| **Methods** | **8** | Specify performance measures, calibration, and utility. | ◀ **Partially reported** | Accuracy, ROC-AUC, F1, sensitivity, specificity reported; calibration unavailable and utility analysis not performed. | **1** |
| **Results** | **9** | Report participant/event numbers. | ✓ **Fully reported** | 1,199 unique patients/lesions: 578 benign, 621 malignant. | **1** |
| **Results** | **10** | Summarize final predictors. | ✓ **Fully reported** | Dermoscopic image features from EfficientNetB0 and Swin-T. | **1** |
| **Results** | **11** | Report estimates with confidence intervals. | ✓ **Fully reported** | Mean accuracy/ROC-AUC, SDs, fold-level 95% CIs, confusion counts, sensitivity, specificity. | **1** |
| **Discussion** | **12** | Interpret main results. | ✓ **Fully reported** | Clinician-facing adjunct; external validation required before clinical use/generalizability claims. | **1** |
| **Registration** | **13** | Provide registration details. | ✓ **Fully reported** | Abstract transparently states study was not registered. | **1** |

### Submission-critical clarifications

**Calibration – partially addressed.** Its importance and absence are reported, but no calibration curve, slope/intercept, or Brier score was produced. It is therefore not represented as a completed quantitative analysis.

**External validation – not performed.** All development and internal validation used the same ISIC-derived cohort with patient- and lesion-independent five-fold partitioning. Independent external validation is required before clinical use or broad generalizability claims.

**Code and reproducibility materials – available on reasonable request.** This includes preprocessing, training, cross-validation, evaluation and interpretability code, model definition, configurations, fold assignments, and checkpoints. No public versioned repository or deployment interface currently exists.

**Data and related materials.** ISIC source images/metadata are publicly available under record-level licences. The filtered cohort manifest, query criteria, associated metadata, and broader ethics-approved protocol are available from the corresponding author on reasonable request, subject to applicable terms.

**Unavailable/not retained.** Extraction/acquisition dates, centre/contributor-specific counts, capture year, device metadata, reason-specific exclusions, original pathology assessor/blinding details, exact hardware/runtime/energy measures, systematic Grad-CAM scoring, and complete image-level audit outputs should remain labelled unavailable unless documentary evidence is recovered.

### Source standards

Collins GS, Moons KGM, Dhiman P, et al. TRIPOD+AI statement: updated guidance for reporting clinical prediction models that use regression or machine learning methods. *BMJ*. 2024;385:e078378. doi:10.1136/bmj-2023-078378.

TRIPOD+AI checklist (11 January 2024) and TRIPOD+AI for Abstracts checklist; medRxiv
